# Heart failure risk is accurately predicted by certain serum proteins

**DOI:** 10.1101/2022.10.11.22280881

**Authors:** V Emilsson, BG Jonsson, V Gudmundsdottir, GT Axelsson, EA Frick, T Jonmundsson, AE Steindorsdottir, LJ Launer, T Aspelund, KA Kortekaas, JH Lindeman, JR Lamb, LL Jennings, V Gudnason

## Abstract

**Aim:** To investigate the utility of serum proteins to predict new-onset heart failure (HF), including those with reduced or preserved ejection fraction (HFrEF or HFpEF), with or without the consideration of known HF-associated clinical variables.

**Methods and results:** The study included 612 participants with HF events from the prospective population-based AGES-Reykjavik cohort of the elderly (N = 5457), 440 of whom were incident cases, with a median follow-up time of 5.45 years. The incident HF population with echocardiographic data included patients with HFrEF (n = 167) and HFpEF (n = 188). The least absolute shrinkage and selection operator (LASSO) model in conjunction with bootstrap resampling validation (500 replications) were used to select predictor variables based on the analysis of 4782 serum proteins and numerous clinical variables related to HF. In at least 80% of bootstrap replications, a subset of 8 to 13 serum proteins had non-zero coefficients for predicting all incident HF, HFpEF, or HFrEF separately. We used C-statistics to assess the goodness of fit when modeling a prognostic risk score for incident HF. In the null model, which did not take age, sex or clinical variables into account, 13 proteins combined had a C-index of 0.80 for all incident HF, whereas for incident HFpEF and HFrEF, the C-index for a subset of 8 or 10 protein predictors combined was 0.78 and 0.80, respectively. The concordance gain for each set of protein predictors was also investigated in the context of the approved biomarker NPPB as well as a number of clinical variables such as Framingham risk score components and calcium in the coronary artery and thoracic aorta. We show that these proteins improve prediction of future HF events even when a large number of HF-associated clinical variables are not included in the model.

**Conclusion:** A small number of circulating proteins were found to accurately predict new-onset HF when no demographic or other information was included, and they also improved the prediction when the main known biomarker NPPB and many HF-associated clinical risk factors of the condition were taken into account.

## Introduction

Heart failure (HF) is a complex clinical syndrome characterized by the heart’s inability to maintain a sufficient blood supply in response to demand. There are many underlying etiologic and pathophysiologic factors that influence the risk of HF. Heart failure is one of the leading causes of morbidity and mortality in developed countries, with a one-year mortality rate of 30% in the US as an example^1^. Moreover, high mortality rates are observed following hospitalization, resulting in readmission reduction as a critical endpoint in recent clinical trials^2^. Heart failure is estimated to have a population prevalence of 2%, which is steadily increasing as the population ages^3^. Approximately equal numbers of newly hospitalized individuals with HF have heart failure with reduced ejection fraction (HFrEF), previously termed systolic heart failure^3^, and heart failure with preserved ejection fraction (HFpEF), previously termed disastolic heart failure^3^.

Recent epidemiological studies revealed that the incidence of acute HFpEF is increasing faster than that of HFrEF^4^, which may be explained by rapid rises in HFpEF associated pathophysiological comorbidities like type 2 diabetes (T2D)^5,6^ and obesity^6,7^. Other comorbidities such as myocardial infarction, uncontrolled hypertension and valve failure are linked to HFrEF^6,8^. It is also known that endothelial dysfunction is more noticeable in HfpEF than in HFrEF, and patients with HFpEF are more likely to be females^6^. In contrast cardiomyocyte loss, fibrosis, and cardiomyocyte hypertrophy are all common symptoms of HFrEF^6^. Heart failure is therefore a heterogenous disorder where the the pathophysiology of the two major subtypes (HFrEF and HFpEF) differ markedly. While there are several pharmacotherapies for HFrEF^9^, no specific treatment for HFpEF is available other than sodium-glucose cotransporter 2 inhibitors (SGLT2)^10^, which are also used to treat HFrEF.

Circulating proteins participate in cross-tissue regulatory loops, which could be a mechanism for system coordination and homeostasis. Similarly, the onset of disease states in individual tissues is most likely the result of an integration of local and global signals^11,12^. In fact, deep serum proteomics has revealed links between circulating proteins and diseases of various etiologies^12-18^, with recent discoveries fueled by aptamer-based affinity methods in particular^13,19-23^. The only approved clinical diagnostic biomarker of HF, N-terminal pro-B-type natriuretic peptide (NPPB)^24^, is readily detected in plasma and serum and is routinely used for HF diagnosis. In the current study, serum levels of 4782 proteins encoded by 4137 human genes from the prospective population-based AGES-RS cohort were tested for their ability to predict future incidence of HF. The capacity of serum proteins to predict HF-related outcomes was examined in the context of multiple clinical variables that have been associated with risk of HF. Using the data-driven least absolute shrinkage and selection operator (LASSO) model in conjunction with bootstrap resampling validations (500 replications), we identified independent serum protein predictors for incident HF, including incident HFpEF and HFrEF, that appear with non-zero estimates in at least 80% of the bootstrap iterations. These proteins were used to construct risk prediction models using C statistics, demonstrating that when they are combined, they accurately predict new-onset HF with or without other information included in the model.

## Methods

### Study population

Cohort participants aged 66 through 96 years at the time of blood collection were from the Age, Gene/Environment Susceptibility Reykjavik Study (AGES-RS)^25^, a single-center prospective population-based study of the elderly (N = 5764, mean age 76.6±6 years) and survivors of the 50-year-long prospective Reykjavik study (N = 19,360). The AGES-RS is an epidemiologic study focusing on four biologic systems: vascular, neurocognitive (including sensory), musculoskeletal, and body composition/metabolism^25^. All AGES-RS participants are of European ancestry. AGES-RS was approved by the National Bioethics Committee in Iceland that acts as the institutional review board for the Icelandic Heart Association (approval number VSN-00-063, in accordance with the Helsinki Declaration) and by the US National Institutes of Health, National Institute on Aging Intramural Institutional Review Board. All participants gave informed, multistage consent prior to enrollment.

The criteria for HF were based on symptoms, signs, chest X-ray, and, in many cases, echocardiographic findings from hospital records, which were adjudicated by examining every record for both prevalent, i.e. HF at the time of recruitment (cross-sectional), and incident HF with a 8-year follow-up from the baseline visit. Individuals with HF were identified by a cardiologist who used hospital discharge ICD-10 codes to determine heart failure outcomes (I50), and this was verified by review of hospital records. Heart failure subtypes were categorized based on echocardiographic criteria. Individuals with HF with preserved ejection fraction (HFpEF) had a left ventricular ejection fraction of 50% or higher, whereas those with HF with reduced ejection fraction (HFrEF) had less than a 40% ejection fraction. Those with mid-range ejection fraction between 40 and 49%, currently called HFmrEF^26^, were not included in the analysis.

Prevalent coronary heart disease (CHD) was defined as previous or prevalent myocardial infarction (MI), coronary artery bypass graft or percutaneous intervention obtained from hospital records at AGES visit. Incident CHD events included fatal CHD or incident non-fatal CHD (International Classification of Diseases (ICD) 9th edition, codes 410, 411, 414, 429, and ICD-10th edition, codes I21–I25), obtained from cause of death registries and hospitalization records from the National University Hospital, the main provider of tertiary care in Iceland. Metabolic syndrome (MetS) is defined by three or more of the following: 1. Fasting glucose ≥ 5.6mmol/L, blood pressure ≥ 140/90 mm Hg, triglycerides ≥ 1.7 mmol/L, high-density lipoprotein (HDL) cholesterol (0< to 0.9 mmol/L males or 0< to 1.0 mmol/L for females), BMI > 30kg/m^2^. Systolic and diastolic blood pressure were measured using a Mercury sphygmomanometer, two-times in a supine position. Hypertension was defined as measured systolic blood pressure of more than 140 mm Hg, diastolic blood pressure of more than 90 mm Hg, self-reported doctor’s diagnosis of hypertension or usage of antihypertensive medications. BMI was calculated as weight (kg) divided by height (in meters) squared, lipoproteins and plasma glucose levels were measured on fasting blood samples. Triglyceride (TG) was measured using enzymatic colorimetry (Roche Triglyceride Assay Kit), HDL cholesterol with an enzymatic in vitro assay (Roche Direct HDL Cholesterol Assay Kit), and glucose was measured using photometry (Roche Hitachi 717 Photometric Analysis System). Low-density lipoprotein (LDL) cholesterol was calculated using the Friedewald equation. Type two diabetes (T2D) was determined from self-reported diabetes, diabetes medication use, or fasting plasma glucose ≥ 7 mmol/L according to ADA guidelines^27^.

The calcium in the coronary arteries (CAC) was quantified using the Agatston scoring method^28^, which was reviewed by four image analysts. Phantom-adjusted CAC was expressed as a sum score for all four coronary arteries, as previously described in greater detail^29^. Calcium in the thoracic aorta (TAC) was quantified using the Agatston method^28^. TAC was scored in the proximal descending thoracic aorta (from the inferior border of the transverse arch to the level of the aortic bulb), and the distal descending thoracic aorta (from the level of the aortic bulb to the bottom of the left ventricular apex). Calcium score of each lesion was calculated by multiplying the lesion area by a density factor derived from the maximum Hounsfield units (HU) within this area.

We assessed survival probability for individual protein predictors in a 12-14-year follow-up study, i.e. both overall survival with 2982 events (all-cause mortality) as well as survival post incident CHD with 692 events. All-cause mortality was determined by the national mortality index with validation performed using death certificates. Follow-up time for overall survival was defined as the time from entry into AGES until death from any cause or end of follow-up (end of year 2016), while follow-up time for survival post incident CHD was defined as the time from 28 days after an incident CHD-event until death from any cause or end of follow-up time.

### Serum protein measurements

Blood samples were collected at the AGES-Reykjavik baseline after an overnight fast, and serum prepared using a standardized protocol and stored in 0.5 mL aliquots at −80°C. Serum protein levels were determined using a multiplex SOMAscan proteomic profiling platform (Novartis V3-5K) which employs SOMAmers (Slow-Off rate Modified Aptamers) that bind to target proteins with high affinity and specificity^12^. The current custom-design SOMAscan platform was built to quantify 5034 protein analytes, 4782 of which measure human proteins from 4137 distinct human genes, in a single serum sample with a focus on proteins that are known or expected to be present extracellularly or on the surface of cells. Of the 5764 AGES-RS participants 5457 were measured for the serum proteome^12^. The aptamer-based proteomics platform measures proteins with femtomole (fM) detection limits and a broad detection range or >8 logs of concentration. To avoid batch or time of processing biases, the order of sample collection and separately, sample processing for protein measurements were randomized and all samples run as a single set at SomaLogic Inc. (Boulder, CO, US). The platform exhibits ∼5% coefficients of variation for median intra- and inter-assay variability^12^. SomaLogic performed the assays in collaboration with Novartis, following the protocol described by our group^12^.

Several metrics, including aptamer specificity through direct tandem mass spectrometry (MS) analysis and inferential assessment via genetic analysis, have been used to determine the performance of the proteomic platform, suggesting strong target specificity throughout the platform^12^. Box-Cox transformation was applied on the protein data, and extreme outlier values excluded, defined as values above the 99.5th percentile of the distribution of 99th percentile cutoffs across all proteins after scaling, resulting in the removal of an average 11 samples per SOMAmer. Previous studies have shown that protein quantitative trait loci (QTLs) were well replicated across different study populations and proteomic technologies^12,22,30^.

### Statistical analysis

The LASSO^31^ model and nonparametric bootstrap^32^ resampling validation were used to approximate the sampling distribution of proteomic variables’ coefficients in age- and sex-adjusted logistic and cox proportional hazards regression models as well as their predictive performance as measured by the area under the curve (AUC) and the Harrel’s concordance index (C-index)^33^. The online supplementary **Appendix S1, Figure S1**, depicts a flowchart of the LASSO and the bootstrap analysis of incident HF. In brief, we created two datasets that were repeated for each bootstrap sample: 1. For the training dataset, the bootstrap selects datapoints from approximately 63% of the samples. These were used to fit the LASSO models and estimate the coefficients. 2. For the testing dataset, the datapoints not selected by the bootstrap and comprising approximately 37% of the data were used to estimate the model’s out-of-sample predictive performance, expressed as AUC and C-index. Following that, we took the training dataset and performed 10-fold cross-validation by dividing the samples into ten equally sized groups and selecting one of the ten groups to exclude from the model fitting process. The LASSO path was then fitted to the remaining nine groups using a set of values for the regularization coefficent’s lambda. The prediction error for the left-out tenth group was calculated for each value of lambda. The prediction errors were averaged for each value of lambda, and each of the ten models fitted in this manner provided an error curve. We use this to determine which value of lambda minimizes the error curve and the LASSO model re-fitted on the entire training dataset, and to return the coefficient values from that model. Finally, the coefficients calculated on the training dataset were used to compute out-of-sample prediction errors on the testing dataset. This procedure was carried out 500 times (online supplementary **Appendix S1, Figure S1**).

A protein was classified as important to the prediction of HF if it had an estimated non-zero coefficient in at least 80% of 500 bootstrap replications. After determining which proteins were important for prediction, we conducted additional analyses to decide whether including these proteins in clinical prediction models would improve prediction quality. This was performed for individual proteins as well as all proteins combined. Here each bootstrap iteration gives us the estimated coefficients for proteins. For the combined protein panel, we used these coefficients to compute a weighted sum of the measured protein values, yielding a single number for each participant that is the log of the hazard ratio for that participant, which we can use to compare the relative hazards between participants. We used the nonparametric bootstrap to approximate the out-of-sample concordance of the models by sequentially adding variables to the models and considering which ones added to the predictive capability by comparing the performance of each model on the same bootstrap samples and calculating 95% confidence intervals on the pairwise differences in AUC and C-index. In all analyses we adjusted for age and sex, and for LASSO models the coefficients for age and sex were unpenalized.

## Results

### Study population and prevalence of risk factors for heart failure

For individuals in the prospective population-based AGES-RS cohort, HF diagnosis was based on hospital records that were adjudicated by examining every record for both prevalent and incident HF with up to eight years of follow-up time (**Methods**). In this cohort, 612 individuals were diagnosed with HF, with 172 diagnosed before the time of blood collection and 440 diagnosed during the follow-up. Of the 612 HF patients with echocardiographic data, 237 had HFrEF and 238 had HFpEF (**Methods**), with 167 and 188 having incident HFrEF and HFpEF, respectively (**Table 1**). The incident cases had a median follow-up time of 5.45 years (range 0.005 to 7.77 years) and an incidence rate of 1.58 cases per 100 person-years at risk. **Table 1** shows selected measures of the AGES-RS cohort including sex stratified demographic, biochemical, clinical, physiological, anthropometric, and imaging baseline characteristics, as well as the prevalence and incidence of HF and HF-related disease endpoints, while **Table 2** compares incident HFpEF to incident HFrEF with regard to the same clinical variables. Online supplementary **Appendix S1, Tables S1**–**S3**, compare all incident HF cases, as well as HFpEF and HFrEF patients separately, to non-HF cases in the AGES-RS cohort.

**Table 1.**
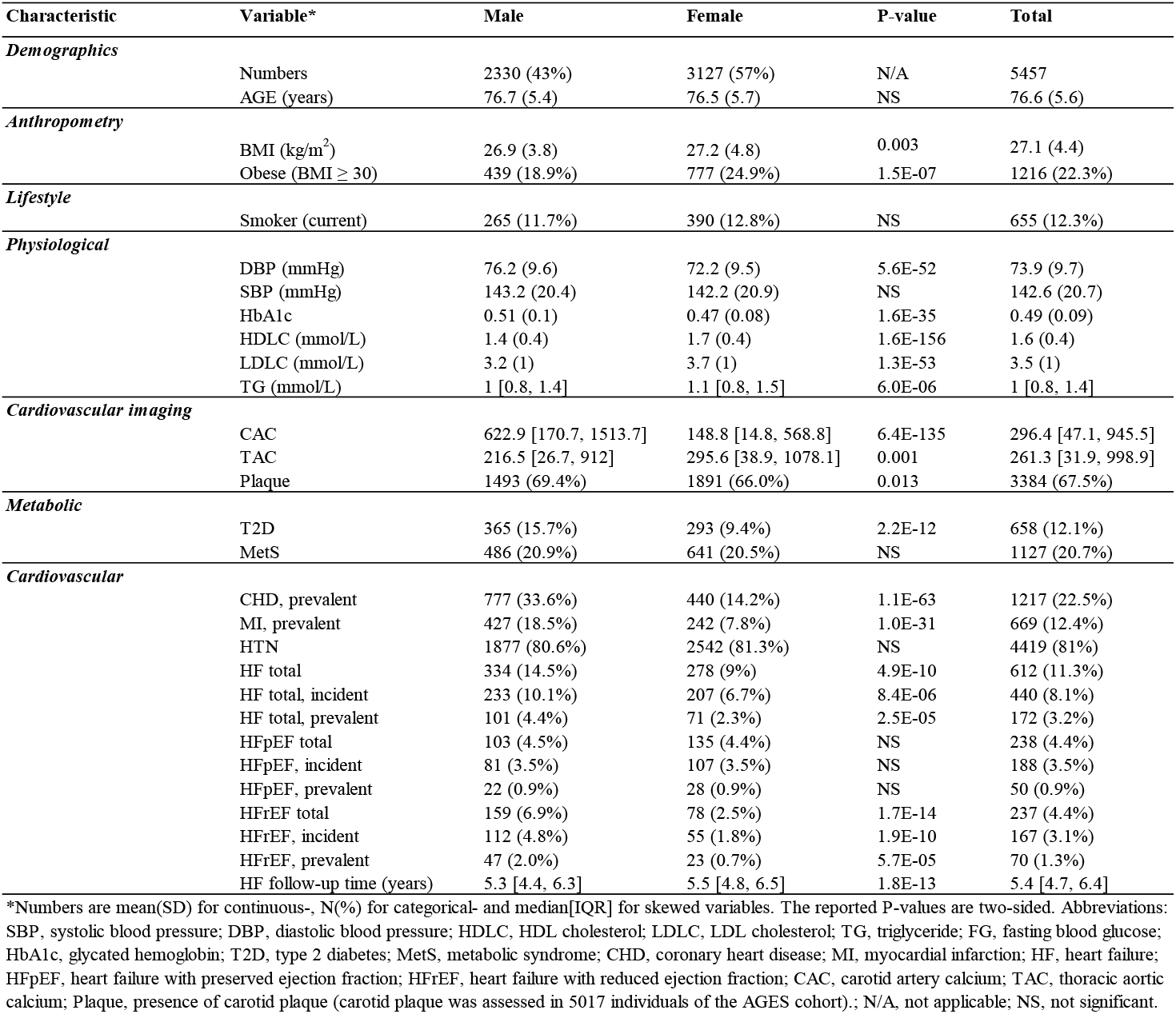
Baseline characteristics of the AGES Reykjavik study cohort.

**Table 2.** Baseline characteristics of incident HFpEF and HFrEF patients in the AGES-RS cohort were compared.

| Characteristic | Variable* | Incident HFpEF | Incident HFrEF | P-value | Total |
| --- | --- | --- | --- | --- | --- |
| **Demographics** |  |  |  |  |  |
|  | Numbers | 188 (53%) | 167 (47%) | N/A | 355 |
|  | Male | 81 (43.1%) | 112 (67.1%) | 9.8E-06 | 193 (54.4%) |
|  | AGE (years) | 79.3 (5.8) | 79.3 (5.6) | NS | 79.3 (5.7) |
| **Anthropometry** |  |  |  |  |  |
|  | BMI (kg/m<sup>2</sup>) | 27.1 (5.3) | 27.5 (4.4) | NS | 27.3 (4.9) |
|  | Obese (BMI ≥ 30) | 48 (25.5%) | 39 (23.4%) | NS | 87 (24.5%) |
| **Lifestyle** |  |  |  |  |  |
|  | Smoker (current) | 28 (15.5%) | 24 (14.6%) | NS | 52 (15.1%) |
| **Physiological** |  |  |  |  |  |
|  | DBP (mmHg) | 72.3 (10.8) | 74.7 (11.2) | 0.0397 | 73.5 (11.1) |
|  | SBP (mmHg) | 147.9 (24.8) | 145.5 (23) | NS | 146.7 (24) |
|  | HbA1c | 0.50 (0.1) | 0.49 (0.09) | NS | 0.49 (0.09) |
|  | HDLc (mmol/L) | 1.6 (0.5) | 1.4 (0.4) | 0.0059 | 1.5 (0.4) |
|  | LDLC (mmol/L) | 3.3 (1) | 3.4 (1.1) | NS | 3.4 (1.1) |
|  | TG (mmol/L) | 1.1 [0.8, 1.5] | 1 [0.8, 1.4] | NS | 1.1 [0.8, 1.5] |
| **Cardiovascular imaging** |  |  |  |  |  |
|  | CAC | 779.2 [182.6, 1502.6] | 974.3 [371.2, 2023.3] | 0.0183 | 869 [265.5, 1683.8] |
|  | TAC | 774.7 [237.4, 2053.4] | 687.7 [174.8, 2175.6] | NS | 752.9 [194.1, 2114] |
|  | Plaque | 143 (83.1%) | 139 (86.3%) | NS | 282 (84.7%) |
| **Metabolic** |  |  |  |  |  |
|  | T2D | 40 (21.3%) | 32 (19.2%) | NS | 72 (20.3%) |
|  | MetS | 52 (27.7%) | 38 (22.8%) | NS | 90 (25.4%) |
| **Cardiovascular** |  |  |  |  |  |
|  | CHD, prevalent | 54 (28.7%) | 73 (43.7%) | 0.0047 | 127 (35.8%) |
|  | MI, prevalent | 21 (11.2%) | 46 (27.5%) | 1.5E-04 | 67 (18.9%) |
|  | HTN | 174 (92.6%) | 155 (92.8%) | NS | 329 (92.7%) |
|  | HF follow-up time (years) | 3.3 [1.9, 4.8] | 3.1 [1.5, 4.5] | NS | 3.2 [1.7, 4.7] |

\*Numbers are mean(SD) for continuous-, N(%) for categorical- and median[IQR] for skewed variables. The reported P-values are two-sided. Abbreviations: SBP, systolic blood pressure; DBP, diastolic blood pressure; HDLC, HDL cholesterol; LDLC, LDL cholesterol; TG, triglyceride; FG, fasting blood glucose; HbA1c, glycated hemoglobin; T2D, type 2 diabetes; MetS, metabolic syndrome; CHD, coronary heart disease; MI, myocardial infarction; HF, heart failure; CAC, carotid artery calcium; TAC, thoracic aortic calcium; Plaque, presence of carotid plaque (carotid plaque was assessed in 5017 individuals of the AGES cohort); N/A, not applicable; NS, not significant.

Males were significantly more common than females among all HF patients, and this was true for both prevalent and incident HF (**Table 1** and online supplementary **Appendix S1, Table S1**), with this difference being most noticeable and only significant among HFrEF patients (**Table 1**, online supplementary **Appendix S1, Tables S2** and **S3**). A direct comparison of HFrEF and HFpEF patient groups (**Table 2**), reveals that 67% of HFrEF patients are males, compared to 43% of HFpEF patients (*P* = 9.6×10^−6^). In fact, the gender proportions of those with HFpEF were comparable to those in the cohort overall, while males with HFrEF far outnumber the population mean (**Table 1** and online supplementary **Appendix S1, Tables S2** and **S3**). It is also clear that males and females in the general AGES-RS cohort differ significantly in terms of many of the known risk factors associated with heart failure (**Table 1**). Males, for example, had significantly higher coronary artery calcium (CAC) than females (CAC score 623 vs. 148, *P* = 6.4×10^−135^), whereas females were more likely to be obese (24.9% vs. 18.9%, *P* = 1.5×10^−7^) (**Table 1**). Furthermore, when comparing the HFrEF and HFpEF patient groups, the HFrEF group was more likely to be affected by MI events and had significantly higher CAC (**Table 2**). This further emphasizes the distinct etiologies of HFrEF and HFpEF. Note also that the incident HF group of both types had significantly higher CAC, thoracic aortic calcium (TAC), and plaque than those who did not have HF (online supplementary **Appendix S1, Tables S2** and **S3**).

### Identifying independent protein predictors of heart failure

In order to identify independent predictors of HF, we estimated the sampling distribution of logistic regression coefficients for all 4782 proteins and a number of known HF risk clinical variables using LASSO regression^31^ and nonparametric bootstrap^32^ resampling for validation (500 replications) (see **Methods**, and online supplementary **Appendix S1, Figure S1**). Online supplementary **Appendix S1, Figure S2**, compares all prevalent and incident HF in terms of multiple clinical variables that may be risk predictors for these conditions. MI and CHD were the most common predictors of prevalent HF among the top clinical variables with non-zero estimates, whereas age, CAC, TAC, and CHD were the most common predictors of incident HF (online supplementary **Appendix S1, Figure S2**).

For serum proteins, most showed no association to HF, with 85% of the 4782 proteins present in less than 10% of the 500 bootstrap replications for prevalent HF (n = 172) and 90% present in fewer than 20% of iterations for incident HF (n = 440), demonstrating that this is a sparse prediction problem. Only proteins selected in at least 80% of the bootstrap replications were considered interesting from this point forward. For prevalent HF, 15 proteins were selected, with renin (REN), parathyroid hormone (PTH), and the only approved clinical diagnostic biomarker of HF, N-terminal pro-B-type natriuretic peptide (NPPB)^24^, always included (online supplementary **Appendix S1, Figure S3A**). In the case of incident HF, 13 proteins were selected, with NPPB always included (online supplementary **Appendix S1, Figure S3B**). Some of these proteins have previously been linked to HF risk in some way, including REN^34^, PTH^35^, alpha-2-HS-glycoprotein (AHSG, aka fetuin A)^36^, troponin I3 cardiac type (TNNI3)^37^, matrix metalloproteinase-12 (MMP12)^38^ and NPPB^39^ (online supplementary **Appendix S1, Figure S3A, B**). Only NPPB was associated with both prevalent and incident HF (online supplementary **Appendix S1, Figure S2A, B**). Online supplementary **Appendix S1, Figure S4A-D**, depicts a receiver operating characteristic curve (ROC) for the diagnostic ability of the protein predictors (with or without NPPB) to classify prevalent or incident HF, demonstrating a significant difference in the area under the curve (AUC) of the ROC curves for demographics versus demographics plus proteins (prevalent HF, AUC 0.65 vs. 0.91, *P* = 4.9×10^−37^; incident HF, AUC 0.68 vs. 0.81, *P* = 1.1×10^−31^). Finally, a sex- and age-adjusted logistic regression analysis of all 4782 proteins (4137 gene symbols) using a study-wide significance threshold (*P* < 1×10^−5^) revealed that many (7 out of 13) of the protein predictors overlap the 318 set of proteins associated with incident HF (online supplementary **Appendix S1, Figure S5A, B**, and online supplementary **Appendix S2**).

Given that our primary interest is in identifying predictors of new-onset HF cases we focused on incident HF in this and subsequent sections. Here, we employed the Harrel’s concordance probability estimate (C-index) to assess the goodness of fit when modeling the prognostic risk scores^33^. **Figure 1** displays the concordance gain for individual proteins, or all proteins combined as a weighted sum of measured coefficients (**Methods**), for predicting incident HF (highlighted in online supplementary **Appendix S1, Figure S3B)**. Concordance gain is demonstrated using several models of adjustment that incorporated various clinical variables and demographic information including the BMI-based Framingham risk score (FRS) components^40^ (age, sex, total cholesterol, HDL cholesterol, systolic blood pressure, smoking status and BMI) plus T2D, and computed tomography (CT) imaging data on coronary artery and thoracic aorta calcification. It is evident that the protein predictors, either with or without NPPB, improve prediction beyond the scope of recognized clinical risk indicators (**Figure 1**). All proteins combined have a C-index of 0.80 in the null model, which includes no demographic or relevant clinical information (**Figure 1**). Individually, NPPB predicted relatively well, as expected, with a C-index of 0.72, but so did a few other proteins, such as TNNI3 and MMP12, which had C-indices of 0.64 and 0.68, respectively (**Figure 1**).

**Figure 1.**
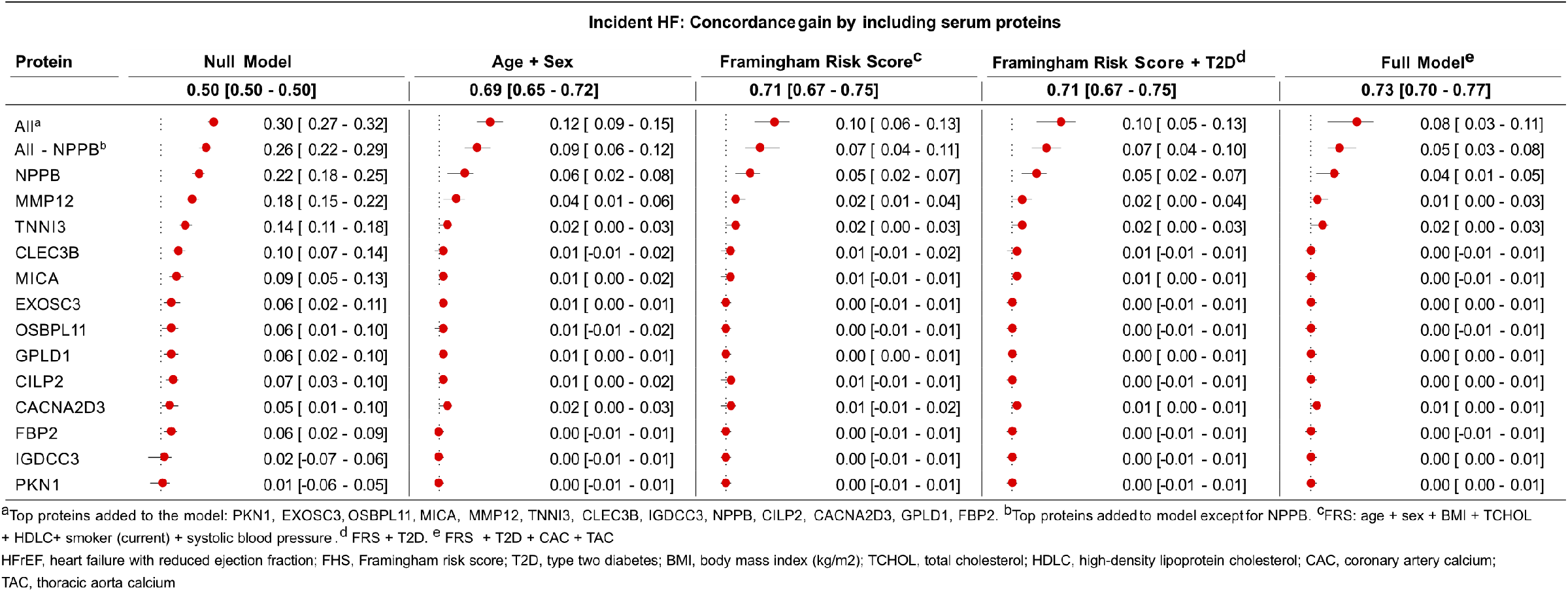
Gain in concordance for serum proteins in predicting, as individual proteins or combined, incident HF. The numbers in each line represent the gain in concordance (C-statistics) over chance in predicting incident HF, with 95% confidence intervals (CIs) in brackets. There is no adjustment for any clinical variable in the null model, but as we move to the right of the figure, we have included an adjustment for age, sex and various clinical variables that are components of the BMI-based FRS, or plus T2D, and in the full adjustment model we have added calcium score of the coronary artery and thoracic aorta. The protein predictors are highlighted individually and collectively in the leftmost column. The C-index for each model for the various clinical variables is shown in bold below each model, with 95% CIs in brackets.

### Serum proteins accurately predict the occurrence of HFrEF and HFpEF

Next, we looked for protein predictors of HFpEF and HFrEF, as well as various clinical variables associated with each condition (see above). **Figure 2A** depicts the best clinical variables that can predict incident HFpEF or HFrEF, whereas **Figure 2B** shows the serum proteins that have non-zero estimates in at least 80% of the bootstrap replications. Age, TAC, and T2D are the key clinical variables in predicting HFpEF, while age, CAC, being male, and having a prior diagnosis of CHD are the most important clinical variables in predicting HFrEF (**Figure 2A**).

**Figure 2.**
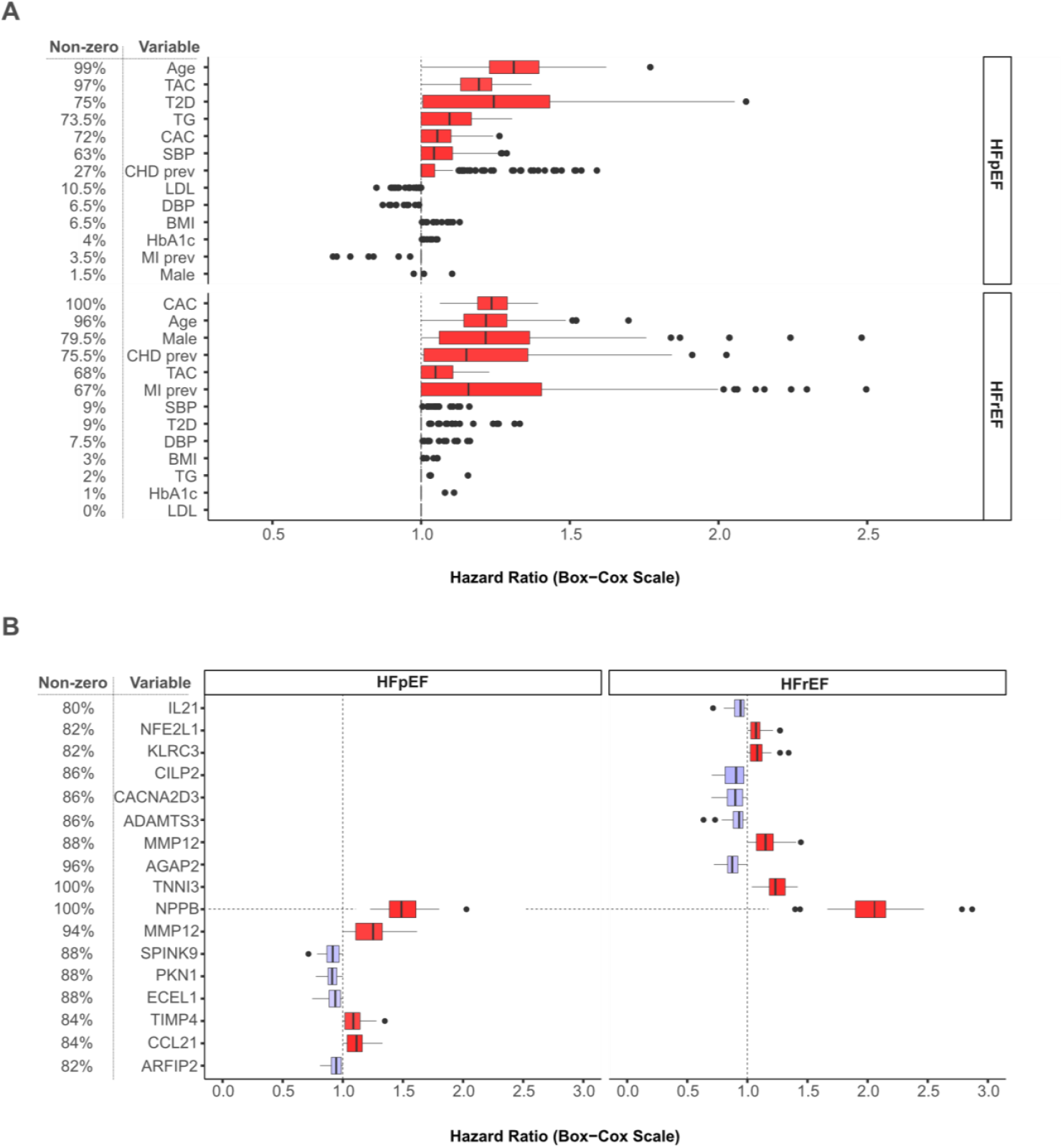
Bootstrap parameter estimates for hazard of HFpEF or HfrEF. **A**. This is shown for clinical variables that are present with non-zero estimates in 80% or more of bootstrap iterations. Abbreviations: TAC, thoracic aorta calcium; T2D, type two diabetes; TG, triglyceride; CAC, coronary artery calcium; **B**. Protein predictors showing up in at least 80% of bootstrap iterations with non-zero estimates for HFpEF or HFrEF.

**Figures 3** and **4** show the concordance gain for the protein predictors of HFpEF and HFrEF, respectively, using several models of adjustment that incorporated various clinical variables based on data from **Figure 2A**. Again, the various protein panels have high prediction scores and also improve prediction beyond known clinical variables associated with HF (**Figures 3** and **4**). In the null model, i.e. no adjustment for clinical variables, all proteins combined have a C-index of 0.78 for HFpEF (**Figure 3**), while the proteins predicting HFrEF have a C-index of 0.80 (**Figure 4**). Interestingly, NPPB and MMP12 predicted HFpEF equally well, both with a C-index of 0.70 (**Figure 3**), whereas metalloproteinase inhibitor 4 (TIMP4) had a relatively good C-index of 0.66. Individual protein predictors such as NPPB, MMP12, and TNNI3 predicted HFrEF with C-indices ranging from 0.67 to 0.74 (**Figure 4**). The ROC curves for the HFrEF and HFpEF protein predictors (with or without NPPB) are shown in online supplementary **Appendix S1, Figure S6A-D**, with a significant difference in the AUC for demographics versus demographics plus proteins (HFrEF, AUC 0.69 vs. 0.81, *P* = 3.2×10^−10^ and HFpEF, AUC 0.64 vs. 0.74, *P* = 7.1×10^−7^). Online supplementary **Appendix S1, Figure S7** depicts the overlap in the number of shared or unique protein predictors for incident HF, HFpEF, and HFrEF. For example, the model selected NPPB and MMP12 for all three incident HF analyses (**Figure 2B** and online supplementary **Appendix S1, Figures S3B** and **S7**). Furthermore, protein kinase N1 (PKN1) was selected for both incident HFpEF and all incident HF, while TNNI3, CILP2, and CACNA2D3 were selected for both incident HFrEF and all incident HF (**Figure 2B** and online supplementary **Appendix S1, Figures S3B** and **S7**). Finally, both HFpEF and HFrEF have a distinct set of five serum protein predictors that are not shared across study groups (**Figure 2B** and online supplementary **Appendix S1, Figures S3B** and **S7**).

**Figure 3.**
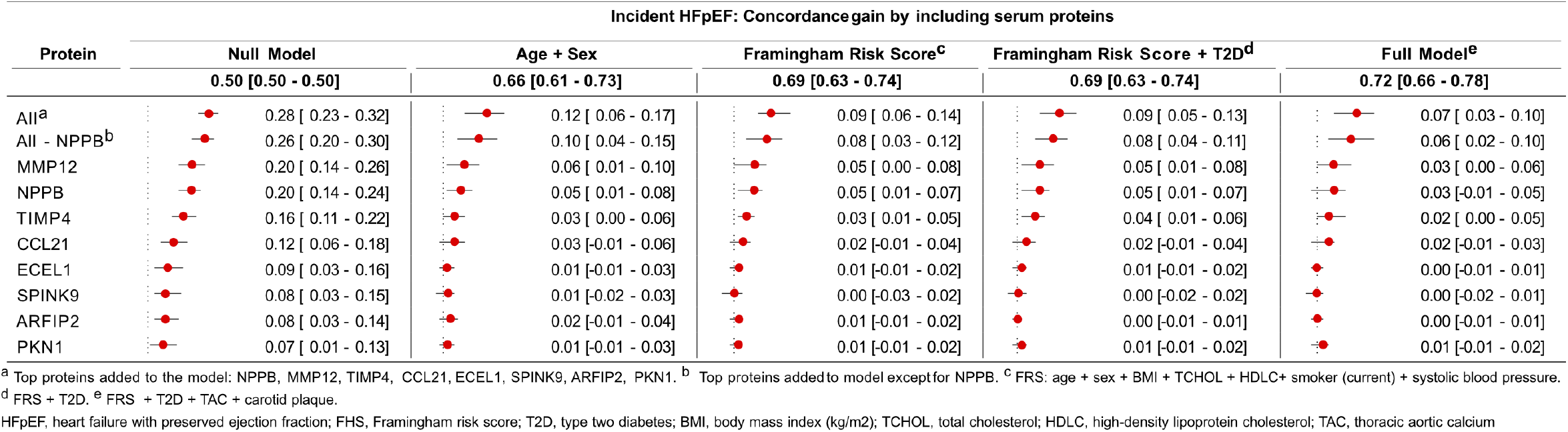
Gain in concordance for serum proteins in predicting, as individual proteins or combined, incident HFpEF. The numbers in each line represent the gain in concordance (C-statistics) over chance in predicting incident HF, with 95% confidence intervals (CIs) in brackets. There is no adjustment for any clinical variable in the null model, but as we move to the right of the figure, we have included an adjustment for age, sex and various clinical variables that are components of the FRS, plus T2D, and the full adjustment model we have added thoracic aortic calcium. The protein predictors are highlighted individually and collectively in the leftmost column. The C-index for each model for the various clinical variables is shown in bold below each model, with 95% CIs in brackets.

**Figure 4.**
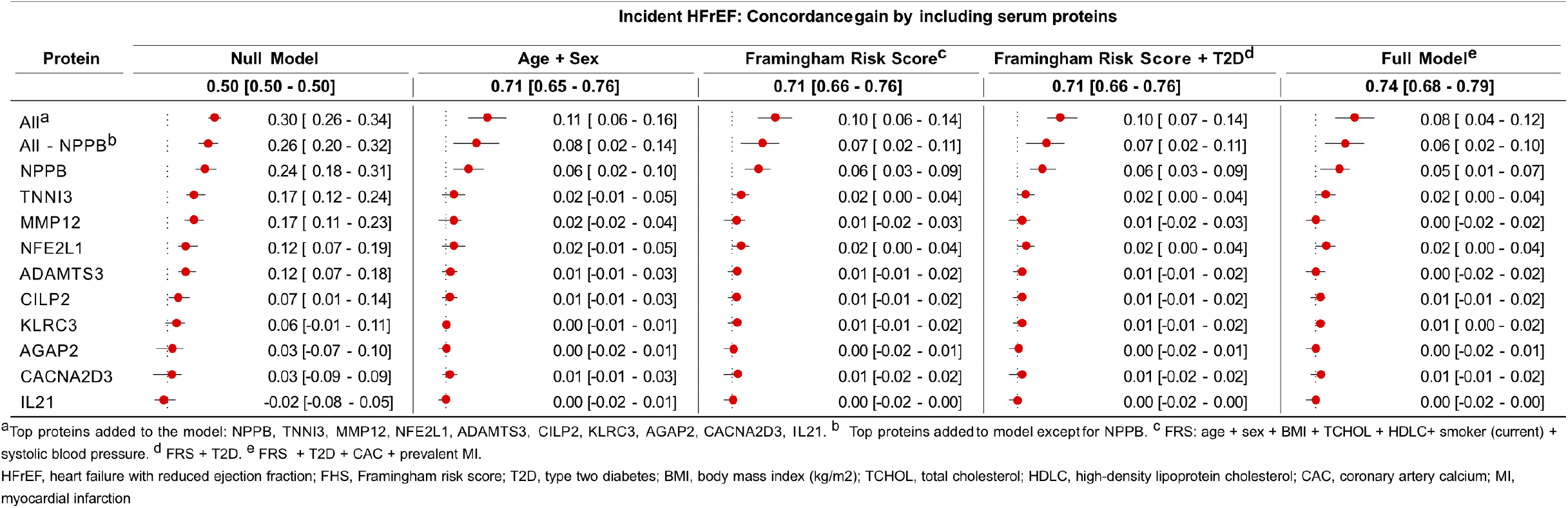
Gain in concordance for serum proteins in predicting, as individual proteins or combined, incident HFrEF. The numbers in each line represent the gain in concordance (C-statistics) over chance in predicting incident HF, with 95% confidence intervals (CIs) in brackets. There is no adjustment for any clinical variable in the null model, but as we move to the right of the figure, we have included an adjustment for age, sex and various clinical variables that are components of the FRS, plus T2D, and the full adjustment model we have added thoracic aortic calcium. The protein predictors are highlighted individually and collectively in the leftmost column. The C-index for each model for the various clinical variables is shown in bold below each model, with 95% CIs in brackets.

Using a sex and age-adjusted logistic regression analysis of all proteins individually and a study-wide significance threshold (*P* < 1×10^−5^), 43 serum proteins (online supplementary **Appendix S1, Figure S8A, B**, and online supplementary **Appendix S2**) were significantly associated with HFpEF, including NPPB, MMP12, and TIMP4 from the panel of protein predictors of HFpEF. A similar analysis linked 12 proteins to HFrEF, including the predictors NPPB, TNNI3, and the nuclear factor erythroid 2-related factor 1 (NFE2L1) (online supplementary **Appendix S1, Figure S9A, B** and online supplementary **Appendix S2**).

**Figures 1, 3**, and **4**, show the C-index for each model for the different clinical variables for different outcomes of incident HF, demonstrating that protein predictor panels alone clearly outperform these models. Inclusion of T2D status had no effect on prediction (**Figures 1, 3** and **4**). Furthermore, **Table 3** compares the C-indices for different models using conventional predictors (age, sex, NPPB, and the FRS) and when the protein predictors for incident HF all, HFpEF, and HFrEF were included. This comparison shows that the newly identified protein predictors improve prediction of well-known HF risk variables. Interestingly, when combined with the protein predictors, the FRS, which includes age, sex, and many clinical variables, does not improve the prediction of new-onset HF compared to using only age and sex in conjunction with the proteins (**Table 3**). The same was true when using the full model, which included FRS and T2D, as well as imaging of calcium in the coronary artery and/or thoracic aorta, carotid plaque, and MI status, depending on the study group (**Table 3**). In other words, compared to using only proteins, including comprehensive information about HF-associated clinical variables added little or nothing to the prediction of new-onset HF.

**Table 3.** C statistics for various models based on HF clinical variables and/or serum proteins. FRS, Framingham risk score components; age, sex, total cholestero HDL cholesterol, systolic blood pressure, smoking status and BMI. Full model: 1. FRS+T2D+TAC+CAC for all incident, 2. FRS+T2D+TAC+carotid p FRS+T2D+CAC+MI

| Model | Incident HF (all) |  |  | Incident HFpEF |  |  | Incident HFrEF |  |  |
| --- | --- | --- | --- | --- | --- | --- | --- | --- | --- |
|  | C-index |  |  | C-index |  |  | C-index |  |  |
|  | Mean | 95% CI lower | 95% CI upper | Mean | 95% CI lower | 95% CI upper | Mean | 95% CI lower | 95% CI upper |
| Age + sex | 0.69 | 0.65 | 0.72 | 0.66 | 0.61 | 0.73 | 0.71 | 0.65 | 0.76 |
| Age + sex + NPPB | 0.74 | 0.71 | 0.78 | 0.72 | 0.67 | 0.76 | 0.77 | 0.70 | 0.81 |
| Age + sex + all proteins | **0.80** | **0.77** | **0.83** | **0.78** | **0.73** | **0.82** | **0.82** | **0.78** | **0.86** |
| FRS | 0.71 | 0.67 | 0.75 | 0.69 | 0.63 | 0.74 | 0.71 | 0.66 | 0.76 |
| FRS + NPPB | 0.76 | 0.73 | 0.80 | 0.73 | 0.68 | 0.78 | 0.77 | 0.72 | 0.83 |
| FRS + all proteins | **0.80** | **0.77** | **0.83** | **0.78** | **0.73** | **0.82** | **0.81** | **0.77** | **0.85** |
| Full model | 0.73 | 0.70 | 0.77 | 0.72 | 0.66 | 0.78 | 0.74 | 0.68 | 0.79 |
| Full model + NPPB | 0.77 | 0.74 | 0.80 | 0.75 | 0.69 | 0.80 | 0.79 | 0.73 | 0.83 |
| Full model + all proteins | **0.81** | **0.78** | **0.84** | **0.78** | **0.73** | **0.83** | **0.82** | **0.78** | **0.86** |

### Characteristics of the proteins that predict and/or are linked to heart failure

This section highlights some of the characteristics of the 23 individual proteins from the different predictors. Online supplementary **Appendix S1, Table S4**, displays the protein predictors’ various properties, such as whether they are secreted, tissue specific, which pathway they are involved in and if there are underlying *cis* and/or *trans*-acting genetic effects in serum. Given the SOMApanel’s design^12^, 37% of the aptamers target secreted proteins, a nearly three-fold enrichment over the 13% of human genes encoding secreted proteins^41^. Intriguingly, 53% of the protein predictors for incident HF are assumed secreted (online supplementary **Appendix S1, Table S4**), a significant enrichment over all secreted proteins targeted by the SOMApanel (F-test *P* = 0.009). Furthermore, seven of the predictor proteins are specifically enriched in the heart, skeletal muscle, or adipose tissue (ECM organization) (online supplementary **Appendix S1, Table S4**). Using gene set collections from the Molecular Signatures Database (MSigDB)^42^ and the GTEx database^43^, we found enrichment in terms related to heart pathology: heart atrial appendage (*P* adj = 0.017) and subcutaneous adipose tissue (*P* adj = 0.038) from GTEX, as well as the KEGG terms dilated cardiomyopathy (*P* adj. = 0.022) and hypertrophic cardiomyopathy (*P* adj. = 0.022) were significantly enriched among our set of 23 proteins. Note also that the broader set of individual proteins that were significantly associated with incident HF in the regression analysis (online supplementary **Appendix S2**, online supplementary **Appendix S1, Figures S5A, S8A**, and **S9A**) are of interest for studies of causal inferences.

Given the numerous clinical variables associated with HF risk examined in the current study, we conducted age- and sex-adjusted linear and logistic regression analyses to see if any of the 23 protein predictors were associated with these traits. The results are summarized in the online supplementary **Appendix S1, Tables S5** and **S6** and **Figure S10**). Further, we included findings from potential associations between these proteins and overall survival probability (all-cause mortality, n = 2982 events) as well as post incident CHD survival probability (n = 692 events) (**Methods**). Some protein predictors, such as MMP12, CILP2, TIMP4 and CLEC3B, were associated with several of these HF-associated clinical traits, whereas others, such as GPLD1, AGAP2, IGDCC3 and IL21 were associated with only a few or none (online supplementary **Appendix S1, Tables S5** and **S6**). It is worth noting that more protein predictors for HFpEF (75% vs. 50%) were linked to metabolic-related traits such as T2D and/or BMI than those for HFrEF (online supplementary **Appendix S1, Tables S5** and **S6**, and **Figure S10**). Furthermore, serum levels of many of these proteins, such as CILP2 and CLEC3B, are directly related to survival probability (online supplementary **Appendix S1, Figure S11**), whereas MMP12 and TIMP4 are inversely related to survival (online supplementary **Appendix S1, Figure S12**).

Two large-scale outcome trials, the DELIVER and EMPEROR studies, recently demonstrated that SGLT2 inhibitors reduce the risk of hospitalization for HF patients with a wide range of ejection fractions, while also improving T2D and kidney function outcomes^10^. In a separate study, the effect of SGLT2 inhibitors on circulating proteins, as measured by the Olink platform (1283 proteins), was investigated in the EMPEROR clinical trial, which included 1134 patients with HFrEF or HFpEF combined^44^. Here, 33 proteins responded to SGLT2 treatment, 25 of which were measured in our study using the aptamer-based technology. Despite the challenge of cross-technology comparison, we found that 14 out of the 25 proteins responding to SGLT2 treatment were significantly associated with incident HF in the current study (online supplementary **Appendix S1, Figure S13A**), 12 of which were significant at the most strict Bonferroni correction for all proteins (*P* < 1×10^−5^), and two (FST and IGFBP1) after correcting for 25 tests (*P* < 0.002). This represents a significant enrichment of proteins responding to SGLT2 treatment among the serum proteins associated with incident HF in our study (OR = 9.4, *P* = 1.6×10^−8^) (online supplementary **Appendix S1, Figure S13B**). For instance, HAVCR1 (aka KIM-1) which was significantly down-regulated in response to 52-week SGLT2 treatment^44^, was positively correlated with incident HF in our study (β = 0.260, *P* = 8.6×10^−7^). The direction of the effect for the other proteins was consistent across studies (online supplementary **Appendix S1, Figure S13A)**. In addition, out of the nine proteins with the largest treatment effect after 12 weeks^44^, five serum proteins, namely IGFBP1, TFRC (aka TfR1), EPO, IGFBP4 and FABP4 (aka AFABP4) (online supplementary **Appendix S1, Figure S13A**), were associated with incident HF. None of the protein predictors were among the proteins that responded to SGLT2 treatment, demonstrating that prediction does not necessitate a causal relationship to outcome^45^.

## Discussion

Heart failure is a chronic, progressive condition with a poor prognosis. It would be a significant advance to develop minimally invasive methods that can predict risk of HF-related hospitalization in the general population. Prior to this study, there were several measures known to be risk factors for HF that could be easily collected (age, sex, BMI), or those generally collected with a cardiologist visit with somewhat greater effort (CAC, TAC, carotid plaque, left ventricular ejection fraction). Currently a combination of many of these characteristics, each measured in a distinct manner is required for the best prediction of HF risk^46^. We investigated the relationship of the above risk factors and the levels of thousands of serum proteins to current (prevalent) and, more importantly, future (incident) cases of HF in the AGES cohort. This resulted in the identification of a small set of proteins that alone, or in combination with known factors more accurately predict HF risk than has been reported to date. Interestingly, the identified proteins appeared to capture the risk associated with various factors (BMI, CAC, plaque, etc.), as well as risk not captured by those factors for new-onset HF, including HFrEF and HFpEF. To put it in other words, measurement of these proteins with a single platform, produced superior results in comparison to measurement of multiple risk factors using multiple platforms. A concordance probability or C-index of between 0.78 and 0.80 was found with proteins alone, even when no demographic or other information was included. Among the protein predictors found in this study was the only approved clinical biomarker of heart failure, NPPB. Importantly, we found that combining the new set of circulating protein predictors with previously approved clinical biomarkers like NPPB and components of the FRS, lead to an improved prediction of new-onset HF.

A total of 23 proteins were selected as predictors by the LASSO model for all incident HF, incident HFpEF, or HFrEF. These proteins were enriched for secretion. It has been proposed that proteins and other biomolecules may be released from solid tissues into blood in response to various cardiac insults, myocardium fibrosis, and inflammatory processes that occur prior to the onset of heart failure^47^. Consistent with this, the protein predictors found here, were enriched in heart and muscle cells, as well as adipose tissue, and were involved in cardiomyopathy-related pathways. A recent study suggests that a complex web of cross-tissue regulatory loops involving serum proteins that connect most or all tissues facilitates systemic homeostasis in humans^11,12^. Similarly, the onset of a syndrome such as HF may also reflect an integration of both local (heart) and global (e.g. adipose tissue) processes that drive the disease state.

There have been some descriptions of plasma protein-based biomarkers linked to HF in the literature, but they are frequently described in a limited case-control context and with few proteins measured. NPPB is the gold standard prognostic biomarker for HF, and it is the only biomarker with a class 1A recommendation for HF diagnosis^24^, so it is not surprising that it appears in all predictor panels of incident HF in our study. Numerous reports of rare monogenic causes of HF, primarily associated with dilated or hypertrophic cardiomyophathies, have been published^48^. In contrast, genome-wide association studies (GWAS) have had limited success in the past in identifying common genetic variations linked specifically to new-onset HFrEF or HFpEF^49^, which is most likely due to the heterogeneous nature of the syndrome’s various forms. However, a recent population based meta-analysis study of 732 incident HF cases using Mendelian randomization (MR) analysis of genetic variants affecting plasma protein levels identified eight proteins, including MMP12, with potential causal relationships to HF^38^. Consistent with this, MMP12 was found to predict both new-onset HFpEF and HFrEF in the current study.

Although there was some clinical variables that predict both HFrEF and HFpEF, others predict one or other subtype: while TAC and T2D are the key clinical variables in predicting HFpEF, being male, CAC and CHD are the clinical variables most likely to predict HFrEF onset. Similarly, while only MMP12 and NPPB were found in all incident protein predictor panels, the other protein predictors for HFrEF versus HFpEF were distinct, which is consistent with these subtypes having different pathobiologies. Other predictors include PKN1 and TIMP4 in the HFpEF predictor panel and TNNI3 in the HFrEF predictor panel. PKN1 deficiency has been linked to systolic and diastolic dysfunction with preserved ejection fraction in a global ischaemia and reperfusion mouse model^50^, whereas pathogenic DNA sequence variants in the TNNI3 are well-established causes of restricted cardiomyopathies^51,52^. The metalloproteinase inhibitor TIMP4 has been linked to heart tissue remodeling and heart failure in rodent models with higher levels of the protein associated with better outcomes^53^. Our data, showed that high serum levels of TIMP4 are associated with an increased risk of HFpEF and lower survival probability, perhaps suggesting a response to underlying pathologies that drive the disease.

Risk prediction models are designed to identify people who are at high risk of developing a disease and can then be targeted for further examination and, eventually, intervention. The predictors do not necessarily imply a causal relationship with the outcome in the sense that modulating a predictor will affect the outcome^45^. Thus, it is not surprising that most of the protein predictors identified were not found among the proteins directly associated with incident HF (online supplementary **Appendix S2**, online supplementary **Appendix 1S, Figures S5, S8** and **S9**). In contrast, those proteins that were differentially expressed in incident HF vs. controls, are likely either causally related to HF or reactive to factors driving HF and as such are potential targets for medical interventions. Similarly, proteins that respond to an HF-drug treatment are likely to be enriched for being causally or reactively linked to the respective outcome. In fact, none of the protein predictors were among the 14 proteins that responded to both SGLT2 treatment and were associated with incident HF (online suplementary **Appendix S1, Figure S13A**). While we anticipate that the direction, i.e. up or down, for proteins responding to SGLT2 treatment to be associated with improved outcome, this may not always be the case in our study’s association with incident HF. In other words, while elevated HAVCR1 levels may be causally related to new-onset HF because they decrease with SGLT2 treatment (online suplementary **Appendix S1, Figure S13A**), the other proteins with a consistent pattern across the two studies are those that may respond to factors that drive the new-onset HF. More generally, however, while signature enrichment is significant, it is difficult to interpret what the direction of change means in functional terms without much more detailed information. So, while HAVCR1 is a great marker of kidney damage and behaves as expected, the others may be causally driving the disease or reacting to it and subject to either positive or negative feedback. Finally, given that nearly four times as many proteins were measured in the current study, these findings suggest that many of the proteins associated with incident HF but not yet investigated for SGLT2 treatment may offer an opportunity to improve our understanding of the biological mechanism underlying the response to HF-drug treatment.

Despite the lack of external validation in this study, it provides a unique opportunity for future protein panel expansion and validation studies for prediction of new-onset HF. Other constraints must be acknowledged. The findings presented were limited to serum proteins and and may not fully capture HF-related pathobiology in solid tissue such as the heart. As all AGES-RS participants are of European ancestry, the results’ transferability and generalizability needs to be tested across all ethnicities. Finally, some of the serum protein predictors for HF in this study are characterized as intracellular proteins, and the significance of their presence in serum, remains to be determined. Our findings lay the groundwork for identifying circulating protein and non-protein biomarkers that can predict future HF, including its subtypes, and as such this could be used in a population surveillance manner outside of the cardiologists office to facilitate early identification of those at risk for HF with either reduced or preserved ejection fraction.

## Conclusion

We used data-driven LASSO analysis in conjunction with bootstrap replications to examine 4782 circulating proteins in serum as well as multiple clinical variables measured in the deeply annotated and aging AGES-RS population-based cohort for prediction of incident HF. A small subset of serum proteins emerged as independent predictors of incident HF, including new-onset HFpEF and HFrEF, complementing previously approved clinical biomarkers like NPPB and the FRS components. These proteins were enriched for secretion, were over-represented in heart and skeletal muscle cells as well as adipose tissue, were enriched in cardiomyopathies-related pathways, and were associated with clinical traits associated with HF as well as survival probability in the AGES-RS. This study offers a unique opportunity for further validation of the protein predictor panel presented, panel expansion with new proteins measured, leading to the development of an accurate and robust prediction panel that, in one platform, in conjunction with currently approved diagnostic tools, could be used by clinical practitioners for early diagnosis and prediction of new-onset HFpEF and HFrEF. The single platform nature of the measures may allow for more extensive at-risk population surveillance by primary care physicians, as well as the possibility of early intervention perhaps in advance of clinical onset of HF.

## Data Availability

The custom-design Novartis SOMAscan data are available through a collaboration agreement with the Novartis Institutes for BioMedical Research. Data from the AGES Reykjavik study are available through collaboration under a data usage agreement with the IHA. All access to data is controlled via the use of a subject-signed informed consent authorization. The time it takes to respond to requests varies depending on their nature and circumstances of the request, but it will not exceed 14 working days. All data supporting the conclusions of the paper are presented in the main text and freely available as a supplement to this manuscript

