## Supporting Information: Appendix S1 for "Heart failure risk is accurately predicted by certain serum proteins"

[1]Icelandic Heart Association, Holtasmari 1, IS-201 Kopavogur, Iceland.
[2]Faculty of Medicine, University of Iceland, 101 Reykjavik, Iceland
[3]Laboratory of Epidemiology and Population Sciences, National Institute on Aging, MD, USA.
[4]Department of Cardiology, Leiden University Medical Center, Leiden, The Netherlands
[5]Department of Surgery, Leiden University Medical Center, Leiden, The Netherlands
[6]Monoceros Biosystems, 12636 High Bluff Drive, Suite 400, San Diego, CA. 92130, USA
[7]Novartis Institutes for Biomedical Research, 22 Windsor Street, Cambridge, MA 02139, USA.

*Shared first authors
[β]Corresponding authors

**Table S1**. Baseline characteristics of HF incident cases in comparison to those without HF, in the AGES-RS cohort.

| Characteristic | Variable* | Incident HF | Without HF | P-value | Total |
|---|---|---|---|---|---|
| ***Demographics*** | | | | | |
| | Numbers | 440 (8%) | 4791 (92%) | N/A | 5231 |
| | Male | 233 (53%) | 1977 (41.3%) | 2.6E-06 | 2210 (42.2%) |
| | AGE (years) | 79.7 (5.7) | 76.3 (5.4) | 1.0E-29 | 76.5 (5.5) |
| ***Anthropometry*** | | | | | |
| | BMI (kg/$m^2$) | 27.3 (5) | 27.0 (4.4) | NS | 27.0 (4.4) |
| | Obese (BMI ≥ 30) | 109 (24.8%) | 1050 (21.9%) | NS | 1159 (22.2%) |
| ***Lifestyle*** | | | | | |
| | Smoker (current) | 66 (15.6%) | 561 (12%) | 0.0392 | 627 (12.3%) |
| ***Physiological*** | | | | | |
| | DBP (mmHg) | 73.1 (10.8) | 74 (9.6) | NS | 74 (9.7) |
| | SBP (mmHg) | 146.7 (24.2) | 142.3 (20.2) | 3.0E-04 | 142.7 (20.6) |
| | HbA1c | 0.50 (0.09) | 0.49 (0.09) | 0.0116 | 0.49 (0.09) |
| | HDLC (mmol/L) | 1.5 (0.5) | 1.6 (0.4) | 7.1E-04 | 1.6 (0.4) |
| | LDLC (mmol/L) | 3.4 (1.1) | 3.5 (1) | 8.3E-03 | 3.5 (1) |
| | TG (mmol/L) | 1.1 [0.8, 1.4] | 1 [0.8, 1.4] | NS | 1 [0.8, 1.4] |
| ***Cardiovascular imaging*** | | | | | |
| | CAC | 837.6 [242.9, 1683.8] | 254.3 [38.2, 830.6] | 3.5E-33 | 289.7 [45.2, 910.1] |
| | TAC | 765.6 [210.5, 2122.4] | 219.6 [26.1, 878.7] | 1.2E-30 | 251.9 [31.1, 962.3] |
| | Plaque | 343 (84.5%) | 2894 (65.6%) | 1.3E-14 | 3237 (67.2%) |
| ***Metabolic*** | | | | | |
| | T2D | 85 (19.3%) | 528 (11%) | 3.4E-07 | 613 (11.7%) |
| | MetS | 108 (24.5%) | 961 (20.1%) | 0.0299 | 1069 (20.4%) |
| ***Cardiovascular*** | | | | | |
| | CHD, prevalent | 162 (36.8%) | 929 (19.4%) | 1.2E-17 | 1091 (20.9%) |
| | MI, prevalent | 88 (20%) | 499 (10.4%) | 1.8E-09 | 587 (11.2%) |
| | HTN | 406 (92.3%) | 3805 (79.4%) | 1.1E-10 | 4211 (80.5%) |
| | HF follow-up time (years) | 3.2 [1.7, 4.6] | 5.5 [4.9, 6.5] | 6.8E-125 | 5.5 [4.7, 6.4] |

*Numbers are mean(SD) for continuous-, N(%) for categorical- and median[IQR] for skewed variables. The reported P-values are two-sided. Abbreviations: SBP, systolic blood pressure; DBP, diastolic blood pressure; HDLC, HDL cholesterol; LDLC, LDL cholesterol; TG, triglyceride; FG, fasting blood glucose; HbA1c, glycated hemoglobin; T2D, type 2 diabetes; MetS, metabolic syndrome; CHD, coronary heart disease; MI, myocardial infarction; HF, heart failure; CAC, carotid artery calcium; TAC, thoracic aortic calcium; Plaque, presence of carotid plaque (carotid plaque was assessed in 5017 individuals of the AGES cohort); N/A, not applicable; NS, not significant.

**Table S2**. Baseline characteristics of incident HFpEF cases in comparison to those without HF, in the AGES-RS cohort.

| Characteristic | Variable* | Incident HFpEF | Without HF | P-value | Total |
|---|---|---|---|---|---|
| ***Demographics*** | | | | | |
| | Numbers | 188 (4%) | 4769 (96%) | N/A | 4957 |
| | Male | 81 (43.1%) | 1967 (41.2%) | NS | 2048 (41.3%) |
| | AGE (years) | 79.3 (5.8) | 76.2 (5.4) | 8.4E-12 | 76.4 (5.5) |
| ***Anthropometry*** | | | | | |
| | BMI (kg/m$^2$) | 27.1 (5.3) | 27 (4.3) | NS | 27 (4.4) |
| | Obese (BMI ≥ 30) | 48 (25.5%) | 1042 (21.9%) | NS | 1090 (22%) |
| ***Lifestyle*** | | | | | |
| | Smoker (current) | 28 (15.5%) | 560 (12%) | NS | 588 (12.2%) |
| ***Physiological*** | | | | | |
| | DBP (mmHg) | 72.3 (10.8) | 74 (9.6) | 0.0330 | 74 (9.6) |
| | SBP (mmHg) | 147.9 (24.8) | 142.3 (20.1) | 0.0027 | 142.5 (20.3) |
| | HbA1c | 0.50 (0.1) | 0.50 (0.09) | NS | 0.50 (0.1) |
| | HDLC (mmol/L) | 1.6 (0.5) | 1.6 (0.4) | NS | 1.6 (0.4) |
| | LDLC (mmol/L) | 3.3 (1) | 3.5 (1) | 0.0221 | 3.5 (1) |
| | TG (mmol/L) | 1.1 [0.8,1.5] | 1 [0.8,1.4] | NS | 1 [0.8,1.4] |
| ***Cardiovascular imaging*** | | | | | |
| | CAC | 779.2 [182.6, 1502.6] | 253.5 [38.2, 826.3] | 5.6E-12 | 267.1 [40.1, 858.2] |
| | TAC | 774.7 [237.4, 2053.4] | 219.6 [25.7, 877.4] | 5.2E-16 | 234.4 [28, 919.4] |
| | Plaque | 143 (83.1%) | 2894 (65.6%) | 2.7E-06 | 3037 (66.3%) |
| ***Metabolic*** | | | | | |
| | T2D | 40 (21.3%) | 523 (11%) | 2.1E-05 | 563 (11.4%) |
| | MetS | 52 (27.7%) | 953 (20%) | 0.0133 | 1005 (20.3%) |
| ***Cardiovascular*** | | | | | |
| | CHD, prevalent | 54 (28.7%) | 923 (19.4%) | 0.0021 | 977 (19.7%) |
| | MI, prevalent | 21 (11.2%) | 496 (10.4%) | NS | 517 (10.4%) |
| | HTN | 174 (92.6%) | 3787 (79.4%) | 1.6E-05 | 3961 (79.9%) |
| | HF follow-up time (years) | 3.3 [1.9, 4.8] | 5.5 [4.9, 6.5] | 6.6E-55 | 5.5 [4.8, 6.5] |

*Numbers are mean(SD) for continuous-, N(%) for categorical- and median[IQR] for skewed variables. The reported P-values are two-sided. Abbreviations: SBP, systolic blood pressure; DBP, diastolic blood pressure; HDLC, HDL cholesterol; LDLC, LDL cholesterol; TG, triglyceride; FG, fasting blood glucose; HbA1c, glycated hemoglobin; T2D, type 2 diabetes; MetS, metabolic syndrome; CHD, coronary heart disease; MI, myocardial infarction; HF, heart failure; CAC, carotid artery calcium; TAC, thoracic aortic calcium; Plaque, presence of carotid plaque (carotid plaque was assessed in 5017 individuals of the AGES cohort); N/A, not applicable; NS, not significant.

**Table S3**. Baseline characteristics of incident HFrEF cases in comparison to those without HF, in the AGES-RS cohort.

| Characteristic | Variable* | Incident HFrEF | Without HF | P-value | Total |
|---|---|---|---|---|---|
| ***Demographics*** | | | | | |
| | Numbers | 167 (3%) | 4769 (97%) | N/A | 4936 |
| | Male | 112 (67.1%) | 1967 (41.2%) | 5.3E-11 | 2079 (42.1%) |
| | AGE (years) | 79.3 (5.6) | 76.2 (5.4) | 1.1E-10 | 76.3 (5.5) |
| ***Anthropometry*** | | | | | |
| | BMI (kg/m$^2$) | 27.5 (4.4) | 27 (4.3) | NS | 27 (4.4) |
| | Obese (BMI ≥ 30) | 39 (23.4%) | 1042 (21.9%) | NS | 1081 (21.9%) |
| ***Lifestyle*** | | | | | |
| | Smoker (current) | 24 (14.6%) | 560 (12%) | NS | 584 (12.1%) |
| ***Physiological*** | | | | | |
| | DBP (mmHg) | 74.7 (11.2) | 74 (9.6) | NS | 74.1 (9.6) |
| | SBP (mmHg) | 145.5 (23) | 142.3 (20.1) | NS | 142.4 (20.2) |
| | HbA1c | 0.50 (0.09) | 0.49 (0.09) | NS | 0.49 (0.09) |
| | HDLC (mmol/L) | 1.4 (0.4) | 1.6 (0.4) | 4.0E-06 | 1.6 (0.4) |
| | LDLC (mmol/L) | 3.4 (1.1) | 3.5 (1) | 0.0221 | 3.5 (1) |
| | TG (mmol/L) | 1 [0.8, 1.4] | 1 [0.8, 1.4] | NS | 1 [0.8, 1.4] |
| ***Cardiovascular imaging*** | | | | | |
| | CAC | 974.3 [371.2, 2023.3] | 253.5 [38.2, 826.3] | 4.4E-21 | 269.3 [40.9, 866.5] |
| | TAC | 687.7 [174.8, 2175.6] | 219.6 [25.7, 877.4] | 1.4E-11 | 229.8 [27.6, 915.9] |
| | Plaque | 139 (86.3%) | 2894 (65.6%) | 7.3E-08 | 3033 (66.3%) |
| ***Metabolic*** | | | | | |
| | T2D | 32 (19.2%) | 523 (11%) | 0.0015 | 555 (11.2%) |
| | MetS | 38 (22.8%) | 953 (20%) | NS | 991 (20.1%) |
| ***Cardiovascular*** | | | | | |
| | CHD, prevalent | 73 (43.7%) | 923 (19.4%) | 2.7E-14 | 996 (20.2%) |
| | MI, prevalent | 46 (27.5%) | 496 (10.4%) | 7.9E-12 | 542 (11%) |
| | HTN | 155 (92.8%) | 3787 (79.4%) | 3.4E-05 | 3942 (79.9%) |
| | HF follow-up time (years) | 3.1 [1.5,4.5] | 5.5 [4.9,6.5] | 1.1E-49 | 5.5 [4.8,6.5] |

*Numbers are mean(SD) for continuous-, N(%) for categorical- and median[IQR] for skewed variables. The reported P-values are two-sided. Abbreviations: SBP, systolic blood pressure; DBP, diastolic blood pressure; HDLC, HDL cholesterol; LDLC, LDL cholesterol; TG, triglyceride; FG, fasting blood glucose; HbA1c, glycated hemoglobin; T2D, type 2 diabetes; MetS, metabolic syndrome; CHD, coronary heart disease; MI, myocardial infarction; HF, heart failure; CAC, carotid artery calcium; TAC, thoracic aortic calcium; Plaque, presence of carotid plaque (carotid plaque was assessed in 5017 individuals of the AGES cohort); N/A, not applicable; NS, not significant.

**Table S4.** Various features of the protein predictors for incident HF (all), incident HFpEF or HFrEF

| Aptamer | Protein predictor | Prediction outcome | Molecular type[a] | Pathway (GO term; BioSystem) | Secreted[b] | Tissue specifcity[c] | Serum cis pQTL[d] | P-value | Serum trans pQTL[d] | P-value |
|---|---|---|---|---|---|---|---|---|---|---|
| 8885-6_3 | CACNA2D3 | incident HF (all), HFrEF | Ion channel | Calcium ion transmembrane transport; dilated cardiomyopathy | No | Non-specific | rs6802227 | 9,00E-18 | rs2301455 | 5,00E-11 |
| 8841-65_3 | CILP2 | incident HF (all), HFrEF | Enzme (protease) | Alkaline phosphatase activity | Yes | Testis | No | N/A | No | N/A |
| 5701-81_3 | CLEC3B | incident HF (all) | Transmembrane $Ca^{2+}$-binding | Skeletal system development | Yes | Adipose tissue (ECM organization) | rs10865936 | 1,00E-29 | No | N/A |
| 12605-1_3 | EXOSC3 | incident HF (all) | Enzyme (hydrolase) | 3'-5'-exoribonuclease activity; metabolism of RNA | No | Non-specific | No | N/A | rs2583341 | 1,00E-08 |
| 9867-23_3 | FBP2 | incident HF (all) | Enzyme (phosphatase) | Fructose metabolic process; metabolic pathways | No | Skeletal muscle | rs10123016 | 3,00E-20 | No | N/A |
| 9500-5_3 | GPLD1 | incident HF (all) | Enzyme (phosphodiesterase) | Ossification; glycosylphosphatidylinositol(GPI)-anchor biosynthesis | Yes | Liver | No | N/A | rs11132382 | 3,00E-18 |
| 7118-24_3 | IGDCC3 | incident HF (all) | Immunoglobulin (membrane) | Neuromuscular process controlling balance | No | Non-specific | No | N/A | No | N/A |
| 2730-58_2 | MICA | incident HF (all) | Glycoprotein | T cell mediated cytotoxicity; | No | Non-specific | rs3128981 | 1E-377 | No | N/A |
| 4496-60_2 | MMP12 | incident HF (all), HFrEF, HFpEF | Enzyme (protease) | Collagen catabolic process | Yes | Non-specific | rs2276109 | 2,00E-125 | No | N/A |
| 7655-11_3 | NPPB | incident HF (all), HFrEF, HFpEF | Hormone | Regulation of vasodilation | Yes | Heart muscle | rs198375 | 5,00E-20 | No | N/A |
| 12878-60_3 | OSBPL11 | incident HF (all) | Lipid-binding | Positive regulation of sequestering of triglyceride | No | Skeletal muscle | rs11575194 | 2,00E-10 | No | N/A |
| 12562-1_3 | PKN1 | incident HF (all), HFpEF | Enzyme (kinase) | Regulation of cell motility; regulation of androgen receptor activity | No | Non-specific | No | N/A | rs12146727 | 3,00E-125 |
| 5441-67_3 | TNNI3 | incident HF (all), HFrEF | Actin-binding | Cardiac muscle contraction; dilated cardiomyopathy | No | Heart left ventricle | No | N/A | rs704 | 4,00E-13 |
| 12630-8_3 | ARFIP2 | HFpEF | Enables protein binding | Phosphatidylinositol-4-phosphate binding; Arf1 pathway | Yes | Non-specific | No | N/A | rs3747207 | 5,00E-12 |
| 2516-57_3 | CCL21 | HFpEF | Chemokine | Establishment of T cell polarity; GPCR ligand binding | Yes | Thyroid | No | N/A | rs2305625 | 1,00E-66 |
| 7076-17_4 | ECEL1 | HFpEF | Enzyme (protease) | Respiratory system process | No | Ovary | No | N/A | No | N/A |
| 8042-88_3 | SPINK9 | HFpEF | Ezyme inhibitor | Serine-type endopeptidase inhibitor activity | Yes | Brain | No | N/A | rs6983956 | 6,00E-13 |
| 6462-12_3 | TIMP4 | HFpEF | Enzyme inhibitor | Negative regulation of metalloenzyme activity | Yes | Adipose tissue (ECM organization) | rs184262 | 6,00E-52 | rs1079734 | 5,00E-30 |
| 8845-2_3 | ADAMTS3 | HFrEF | Enzyme (protease) | Metalloendopeptidase activity and collagen fibril organization | Yes | Retina | No | N/A | No | N/A |
| 14291-53_3 | AGAP2 | HFrEF | Enzyme inhibitor | GTPase activator activity; Netrin-1 signaling;Endocytosis | No | Brain | No | N/A | No | N/A |
| 7124-18_3 | IL21 | HFrEF | Cytokine | Positive regulation of cytokine production; cytokine-cytokine receptor interaction | Yes | Lymphoid tissue | No | N/A | rs78593564 | 1,00E-49 |
| 7795-14_3 | KLRC3 | HFrEF | Transmembrane receptor | Cellular defense response; antigen processing and presentation | No | Non-specific | No | N/A | rs112689088 rs497239 | 1E-248 1E-31 |
| 11154-3_3 | NFE2L1 | HFrEF | Tracscription regulator | RNA polymerase II distal enhancer sequence-specific DNA binding; transcription Regulator | Yes | Skeletal muscle | No | N/A | No | N/A |

[a]Protein classes were manually curated based on information from the Gene Ontology (GO) and Swiss-Prot databases

[b]Derived from Uhlen et al. (PMID: 25613900).

[c]Tissue distribution of RNA and/or protein expression for each serum protein predictor using the GTEx (PMID: 32913098) and The Human Protein Atlas (PMID: 25613900) databases, respectively.

[d]Derived from Gudjonsson et al. (PMID: 35078996).

**Table S5.** Age- and sex-adjusted logistic regression analysis of the association of the protein predictors to various outcomes in the AGES-RS

| Aptamer | Protein target | Prediction outcome | T2D | | MI | | CHD | | Plaque | | Smoker (current) | | Survival (all-cause mortality) | | Survival (post incident CHD) | |
|---|---|---|---|---|---|---|---|---|---|---|---|---|---|---|---|---|
| | | | beta | P-value | beta | P-value | beta | P-value | beta | P-value | beta | P-value | HR | P-value | HR | P-value |
| 8885-6_3 | CACNA2D3 | Incident HF (all), HFrEF | -0,223 | 9,0E-08 | N/A | NS | N/A | NS | N/A | NS | -0,480 | 1,3E-29 | 0,862 | 6,1E-15 | 0,902 | 0,006 |
| 8841-65_3 | CILP2 | Incident HF (all), HFrEF | -0,607 | 2,2E-46 | N/A | NS | N/A | NS | -0,100 | 0,001 | -0,481 | 3,0E-30 | 0,827 | <2E-16 | 0,882 | 0,001 |
| 5701-81_3 | CLEC3B | Incident HF (all) | -0,473 | 1,4E-28 | -0,155 | 0,0003 | -0,194 | 2E-08 | -0,119 | 0,0002 | -0,209 | 1,1E-06 | 0,800 | <2E-16 | 0,820 | 5,1E-07 |
| 12605-1_3 | EXOSC3 | Incident HF (all) | N/A | NS | N/A | NS | N/A | NS | N/A | NS | -0,137 | 0,001 | N/A | NS | N/A | NS |
| 9867-23_3 | FBP2 | Incident HF (all) | N/A | NS | N/A | NS | N/A | NS | N/A | NS | -0,267 | 1,6E-09 | N/A | NS | N/A | NS |
| 9500-5_3 | GPLD1 | Incident HF (all) | N/A | NS | N/A | NS | N/A | NS | N/A | NS | N/A | NS | N/A | NS | N/A | NS |
| 7118-24_3 | IGDCC3 | Incident HF (all) | N/A | NS | N/A | NS | N/A | NS | N/A | NS | N/A | NS | N/A | NS | N/A | NS |
| 2730-58_2 | MICA | Incident HF (all) | N/A | NS | N/A | NS | 0,155 | 6,1E-06 | N/A | NS | N/A | NS | N/A | NS | 1,068 | 0,0004 |
| 4496-60_2 | MMP12 | Incident HF (all), HFrEF, HFpEF | 0,232 | 2,1E-07 | 0,358 | 3,7E-15 | 0,412 | 9,4E-29 | 0,340 | 3,4E-24 | 0,532 | 4,1E-30 | 1,263 | <2E-16 | 1,244 | 2,2E-07 |
| 7655-11_3 | NPPB | Incident HF (all), HFrEF, HFpEF | N/A | NS | 0,618 | 2,2E-39 | 0,656 | 8,6E-65 | 0,188 | 2,2E-08 | N/A | NS | 1,334 | <2E-16 | 1,314 | 9,4E-11 |
| 12878-60_3 | OSBPL11 | Incident HF (all) | N/A | NS | N/A | NS | N/A | NS | N/A | NS | N/A | NS | N/A | NS | N/A | NS |
| 12562-1_3 | PKN1 | Incident HF (all), HFpEF | -0,141 | 0,0008 | N/A | NS | N/A | NS | N/A | NS | N/A | NS | N/A | NS | N/A | NS |
| 5441-67_3 | TNNI3 | Incident HF (all), HFrEF | N/A | NS | 0,167 | 4,9E-05 | N/A | NS | N/A | NS | N/A | NS | 1,107 | 3,4E-08 | N/A | NS |
| 12630-8_3 | ARFIP2 | Incident HFpEF | -0,734 | 9,6E-63 | N/A | NS | N/A | NS | N/A | NS | N/A | NS | 0,900 | 1,1E-08 | 0,892 | 0,001 |
| 2516-57_3 | CCL21 | Incident HFpEF | 0,165 | 0,0001 | N/A | NS | 0,131 | 0,0002 | N/A | NS | N/A | NS | 1,081 | 5,5E-05 | N/A | NS |
| 7076-17_4 | ECEL1 | Incident HFpEF | N/A | NS | N/A | NS | N/A | NS | N/A | NS | N/A | NS | 0,935 | 0,0003 | N/A | NS |
| 8042-88_3 | SPINK9 | Incident HFpEF | -0,291 | 6,2E-12 | N/A | NS | N/A | NS | N/A | NS | 0,207 | 1,5E-06 | 0,865 | 1,9E-14 | 0,885 | 0,001 |
| 6462-12_3 | TIMP4 | Incident HFpEF | N/A | NS | N/A | NS | N/A | NS | 0,110 | 0,0008 | N/A | NS | 1,241 | <2E-16 | 1,176 | 3,1E-05 |
| 8845-2_3 | ADAMTS3 | Incident HFrEF | -0,200 | 2,3E-06 | N/A | NS | -0,111 | 0,001 | N/A | NS | -0,204 | 2,9E-06 | 0,880 | 1,9E-11 | N/A | NS |
| 14291-53_3 | AGAP2 | Incident HFrEF | N/A | NS | N/A | NS | N/A | NS | N/A | NS | N/A | NS | N/A | NS | N/A | NS |
| 7124-18_3 | IL21 | Incident HFrEF | N/A | NS | N/A | NS | N/A | NS | N/A | NS | N/A | NS | N/A | NS | N/A | NS |
| 7795-14_3 | KLRC3 | Incident HFrEF | N/A | NS | 0,177 | 4,2E-05 | 0,117 | 0,0007 | N/A | NS | N/A | NS | 1,119 | 7,6E-09 | N/A | NS |
| 11154-3_3 | NFE2L1 | Incident HFrEF | N/A | NS | N/A | NS | N/A | NS | N/A | NS | N/A | NS | N/A | NS | N/A | NS |

**Table S6.** Age- and sex-adjusted linear regression analysis of the association of the protein predictors to various outcomes in the AGES-RS

| Aptamer | Protein target | Prediction outcome | TCHOL | | LDLC | | HDLC | | TG | | HbA1c | | BMI | | SBP | | DBP | | CAC | | TAC | |
|---|---|---|---|---|---|---|---|---|---|---|---|---|---|---|---|---|---|---|---|---|---|---|
| | | | beta | P-value | beta | P-value | beta | P-value | beta | P-value | beta | P-value | beta | P-value | beta | P-value | beta | P-value | beta | P-value | beta | P-value |
| 8885-6_3 | CACNA2D3 | Incident HF (all), HFrEF | N/A | NS | N/A | NS | 0,067 | 2,6E-30 | -0,114 | 1,6E-37 | -0,006 | 2,8E-06 | -0,772 | 4,7E-37 | N/A | NS | N/A | NS | N/A | NS | N/A | NS |
| 8841-65_3 | CILP2 | Incident HF (all), HFrEF | 0,048 | 0,002 | 0,050 | 0,0004 | 0,033 | 1,8E-08 | -0,075 | 3,6E-17 | -0,021 | 7,9E-60 | N/A | NS | N/A | NS | N/A | NS | N/A | NS | -69,508 | 0,0004 |
| 5701-81_3 | CLEC3B | Incident HF (all) | N/A | NS | N/A | NS | 0,078 | 8,3E-30 | -0,167 | 1,2E-56 | -0,017 | 1,5E-25 | -0,333 | 4,5E-06 | N/A | NS | N/A | NS | -80,361 | 1,8E-06 | -129,289 | 4,9E-08 |
| 12605-1_3 | EXOSC3 | Incident HF (all) | N/A | NS | N/A | NS | N/A | NS | -0,038 | 1,3E-05 | N/A | NS | N/A | NS | N/A | NS | N/A | NS | N/A | NS | N/A | NS |
| 9867-23_3 | FBP2 | Incident HF (all) | N/A | NS | N/A | NS | 0,021 | 0,0003 | N/A | NS | -0,005 | 7,3E-05 | N/A | NS | N/A | NS | N/A | NS | N/A | NS | N/A | NS |
| 9500-5_3 | GPLD1 | Incident HF (all) | N/A | NS | N/A | NS | N/A | NS | N/A | NS | N/A | NS | N/A | NS | N/A | NS | N/A | NS | N/A | NS | N/A | NS |
| 7118-24_3 | IGDCC3 | Incident HF (all) | N/A | NS | 0,045 | 0,001 | N/A | NS | N/A | NS | N/A | NS | N/A | NS | N/A | NS | N/A | NS | N/A | NS | N/A | NS |
| 2730-58_2 | MICA | Incident HF (all) | -0,072 | 2,7E-06 | -0,070 | 9,5E-07 | N/A | NS | N/A | NS | N/A | NS | 0,242 | 8,1E-05 | N/A | NS | N/A | NS | 44,315 | 0,002 | N/A | NS |
| 4496-60_2 | MMP12 | Incident HF (all), HFrEF, HFpEF | -0,065 | 3,5E-05 | N/A | NS | -0,081 | 1,2E-41 | 0,076 | 3,3E-16 | N/A | NS | N/A | NS | N/A | NS | -0,799 | 4,1E-09 | 179,593 | 6,2E-35 | 235,080 | 2,1E-30 |
| 7655-11_3 | NPPB | Incident HF (all), HFrEF, HFpEF | -0,261 | 3,8E-61 | -0,226 | 2,8E-53 | -0,006 | NS | -0,065 | 6,0E-12 | -0,011 | 1,3E-15 | -0,282 | 1,0E-05 | 2,468 | 1,2E-16 | -0,430 | 0,002 | 187,650 | 6,7E-37 | 257,992 | 2,6E-35 |
| 12878-60_3 | OSBPL11 | Incident HF (all) | -0,048 | 0,001 | N/A | NS | -0,018 | 0,002 | N/A | NS | N/A | NS | N/A | NS | N/A | NS | N/A | NS | N/A | NS | N/A | NS |
| 12562-1_3 | PKN1 | Incident HF (all), HFpEF | N/A | NS | -0,047 | 0,001 | N/A | NS | N/A | NS | N/A | NS | -0,210 | 0,0007 | N/A | NS | N/A | NS | N/A | NS | N/A | NS |
| 5441-67_3 | TNNI3 | Incident HF (all), HFrEF | N/A | NS | N/A | NS | 0,031 | 2,0E-07 | -0,043 | 2,4E-06 | N/A | NS | N/A | NS | 0,942 | 0,001 | N/A | NS | 74,078 | 3,0E-07 | 62,892 | 0,002 |
| 12630-8_3 | ARFIP2 | Incident HFpEF | 0,089 | 6,7E-09 | 0,068 | 1,9E-06 | 0,071 | 1,1E-33 | -0,107 | 6,0E-33 | -0,020 | 4,0E-49 | -0,876 | 4,2E-47 | N/A | NS | N/A | NS | -77,029 | 5,9E-08 | N/A | NS |
| 2516-57_3 | CCL21 | Incident HFpEF | -0,127 | 1,2E-16 | -0,073 | 2,6E-07 | -0,055 | 1,3E-20 | N/A | NS | N/A | NS | 0,383 | 4,2E-10 | N/A | NS | N/A | NS | 112,268 | 2,8E-15 | 65,074 | 0,001 |
| 7076-17_4 | ECEL1 | Incident HFpEF | 0,068 | 0,0002 | 0,057 | 0,0007 | 0,033 | 1,3E-06 | -0,047 | 8,4E-06 | N/A | NS | -0,307 | 2,1E-05 | N/A | NS | N/A | NS | N/A | NS | N/A | NS |
| 8042-88_3 | SPINK9 | Incident HFpEF | N/A | NS | N/A | NS | 0,054 | 3,8E-21 | -0,079 | 1,7E-19 | -0,006 | 1,1E-05 | -0,805 | 7,4E-42 | N/A | NS | -0,479 | 0,0002 | N/A | NS | N/A | NS |
| 6462-12_3 | TIMP4 | Incident HFpEF | -0,058 | 0,0003 | -0,075 | 2,9E-07 | 0,056 | 1,7E-20 | -0,087 | 4,1E-21 | -0,012 | 9,3E-17 | N/A | NS | N/A | NS | N/A | NS | 96,642 | 3,9E-11 | 132,126 | 1,4E-10 |
| 8845-2_3 | ADAMTS3 | Incident HFrEF | 0,057 | 0,0003 | 0,052 | 0,0003 | 0,034 | 1,6E-08 | -0,065 | 2,2E-12 | -0,008 | 9,4E-09 | N/A | NS | N/A | NS | N/A | NS | N/A | NS | N/A | NS |
| 14291-53_3 | AGAP2 | Incident HFrEF | N/A | NS | N/A | NS | N/A | NS | N/A | NS | N/A | NS | N/A | NS | N/A | NS | N/A | NS | N/A | NS | N/A | NS |
| 7124-18_3 | IL21 | Incident HFrEF | N/A | NS | N/A | NS | N/A | NS | N/A | NS | N/A | NS | N/A | NS | N/A | NS | N/A | NS | N/A | NS | N/A | NS |
| 7795-14_3 | KLRC3 | Incident HFrEF | N/A | NS | -0,049 | 0,0008 | -0,028 | 4,4E-06 | 0,076 | 2,4E-16 | -0,005 | 0,0002 | 0,328 | 1,8E-07 | N/A | NS | N/A | NS | 89,704 | 9,7E-10 | 64,098 | 0,002 |
| 11154-3_3 | NFE2L1 | Incident HFrEF | -0,049 | 0,001 | N/A | NS | -0,021 | 0,0003 | N/A | NS | N/A | NS | N/A | NS | N/A | NS | N/A | NS | N/A | NS | N/A | NS |

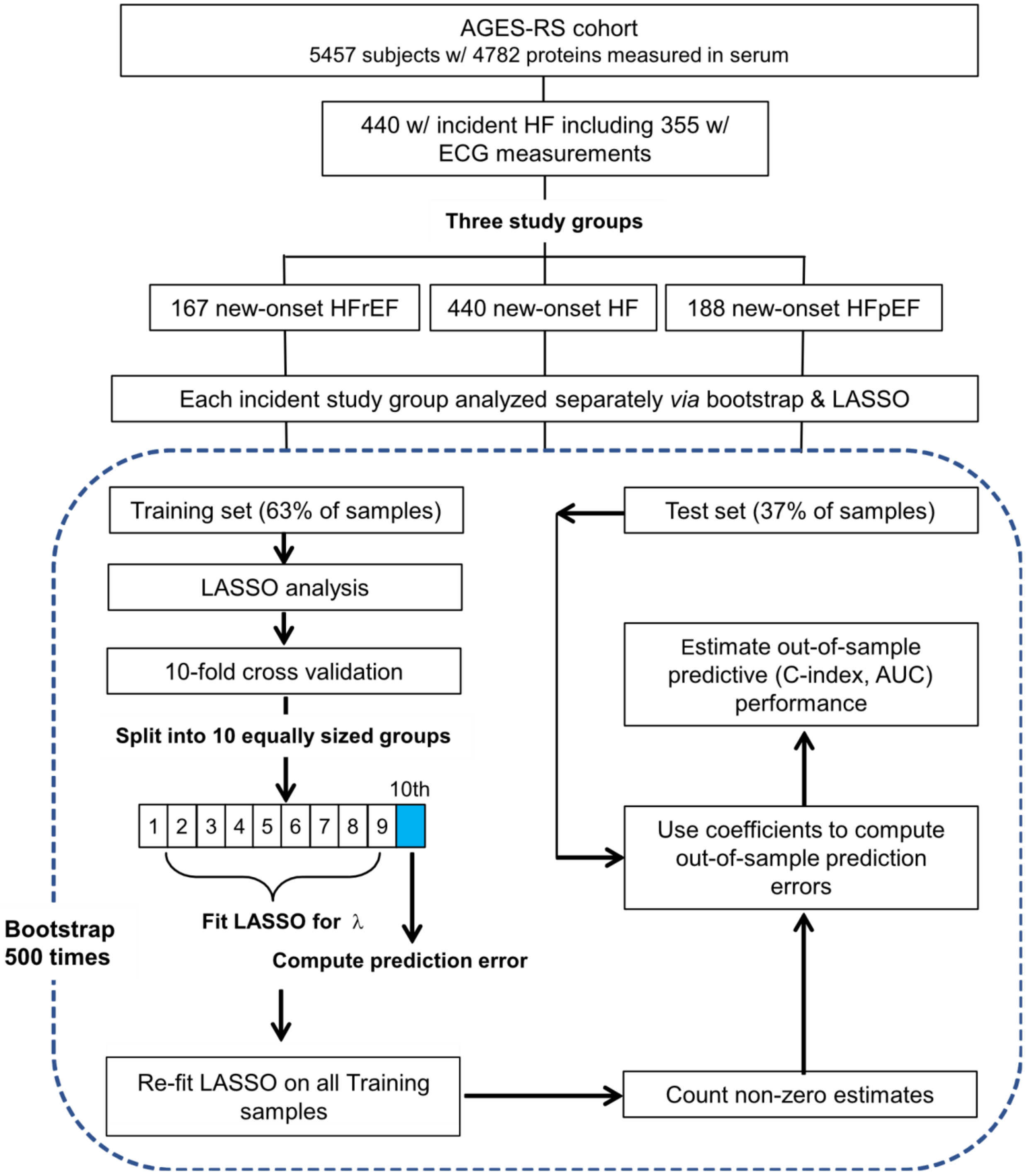


**Figure S1. Flowchart of the current study's bootstrap and LASSO analyses**. The Method section contains a more detailed description of the bootstrap and LASSO analyses used to generate the best performing predictors (C-index).

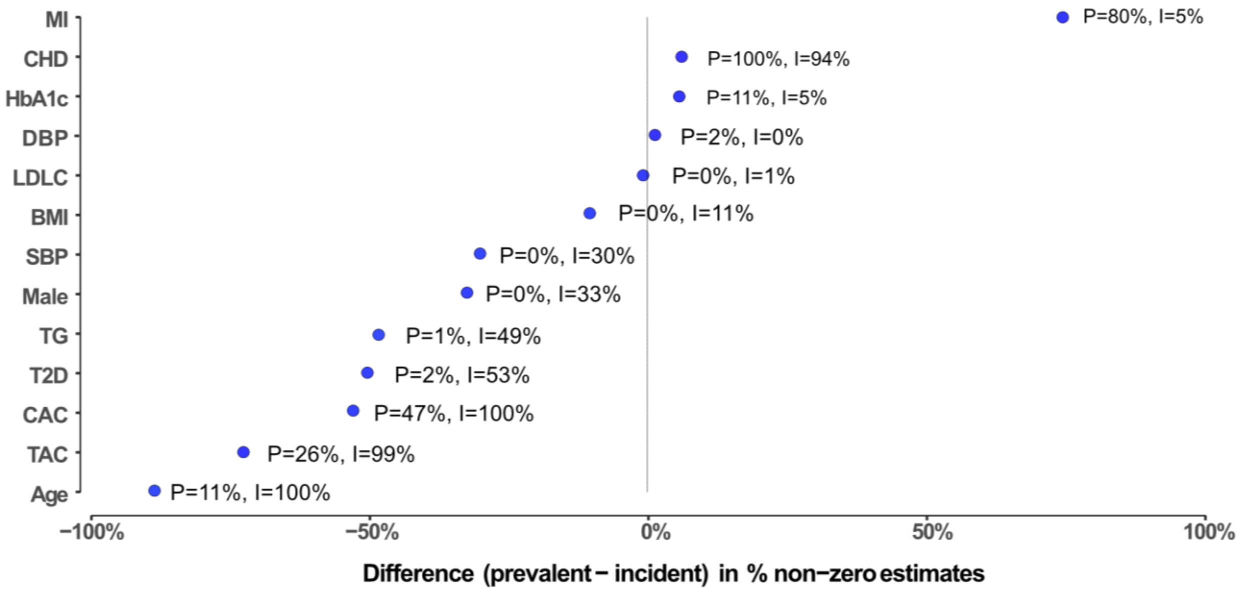


**Figure S2**. **Comparing prevalent and incident HF in terms of clinical variables as clinical predictors**. The x-axis represents the difference in percentage non-zero estimates from 500 bootstrap iterations combined with LASSO regression between prevalent (P) and incident (I) HF, while the y-axis represents clinical variables as predictors. Abbreviations: MI, myocardial infarction; CHD, coronary heart disease; HbA1c; glycosylated hemoglobin; DBP, diastolic blood pressure; LDLC, low-density lipoprotein cholesterol; BMI, body mass index; SBP, systolic blood pressure; TG, triglyceride; T2D, type two diabetes; CAC, coronary calcium; TAC, thoracic aorta calcium.

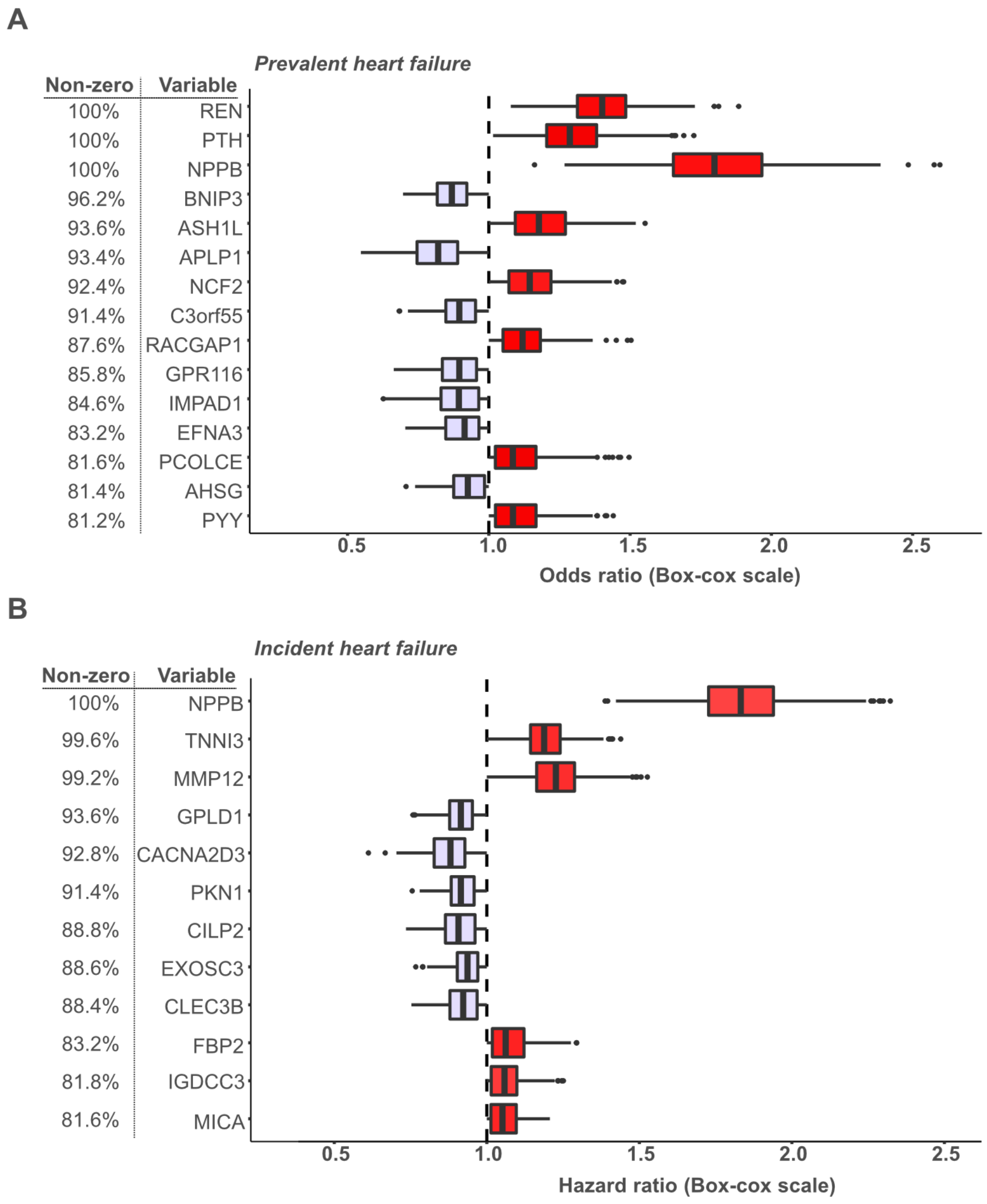


**Figure S3**. **Estimates of bootstrap parameters for serum proteins predicting the risk of prevalent or incident HF**. **A**. the figure depicts protein predictors for prevalent HF using a data-driven nonparametric bootstrap and LASSO regression analysis (see Methods for more detail). On the left, we show the percentage of iterations with non-zero coefficients for the corresponding protein variables that appear in at least 80% of iterations, while the x-axis shows the mean estimates (odds ratio) and 95% confidence intervals. **B**. a similar plot highlighting the protein predictors for incident HF with the estimates (x-axis) as the hazard ratio.

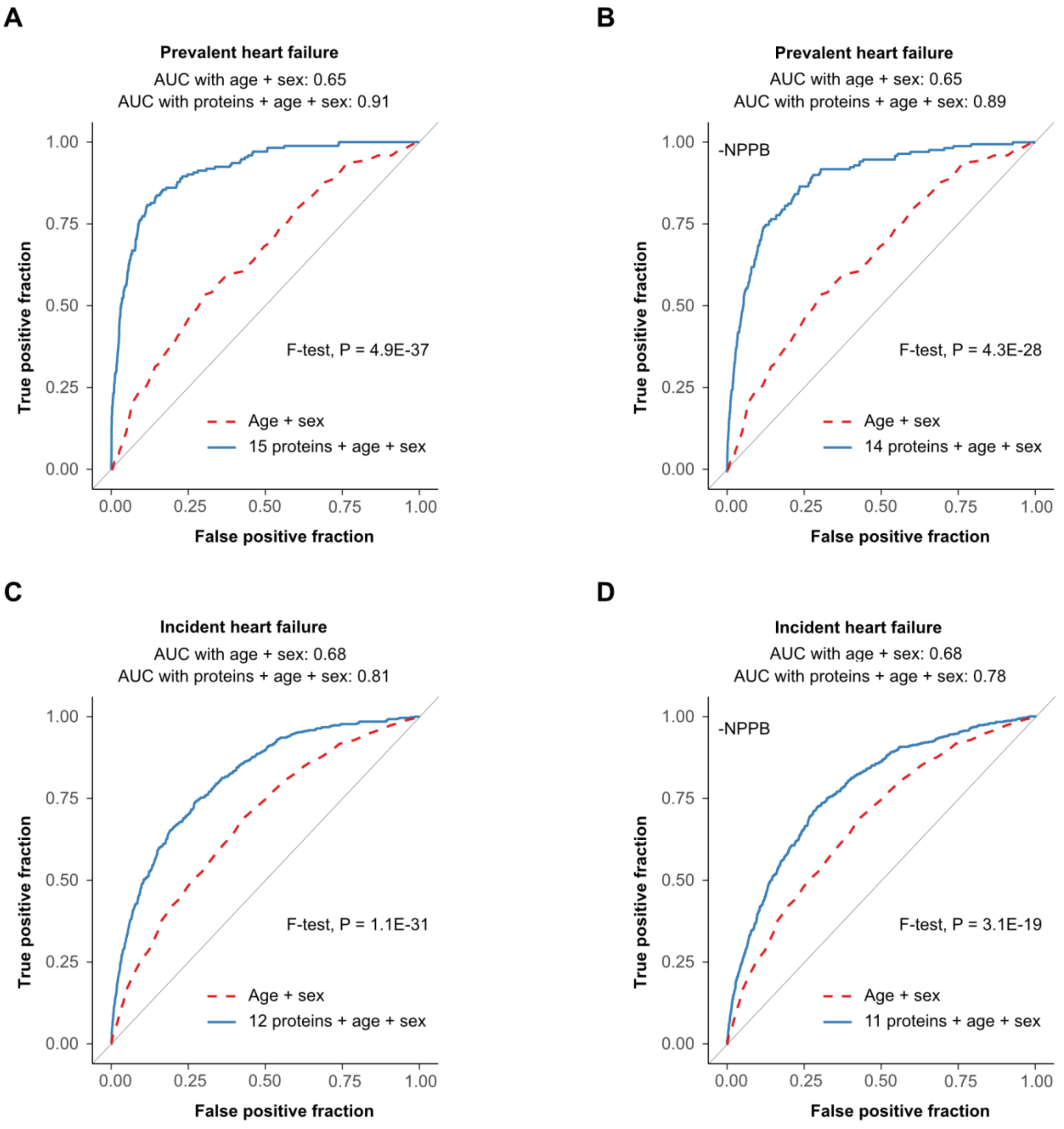


**Figure S4**. **The ability of serum protein predictors to classify prevalent or incident HF**. A receiver operating characteristic curve (ROC) for the ability of **A**. all 15 protein predictors from the online supplementary Figure S2A to classify prevalent HF and B. when NPPB is excluded from the protein panel. C. depicts the ROC curve for all protein predictors from the online supplementary Figure SB for incident HF, while D. excludes NPPB from the classifier. The F-tests of equality P-values (two-sided) show significant differences between the demographics (age + sex, red broken curve) ROC curves and the demographics plus proteins (age + sex + proteins, blue solid curve).

A

B

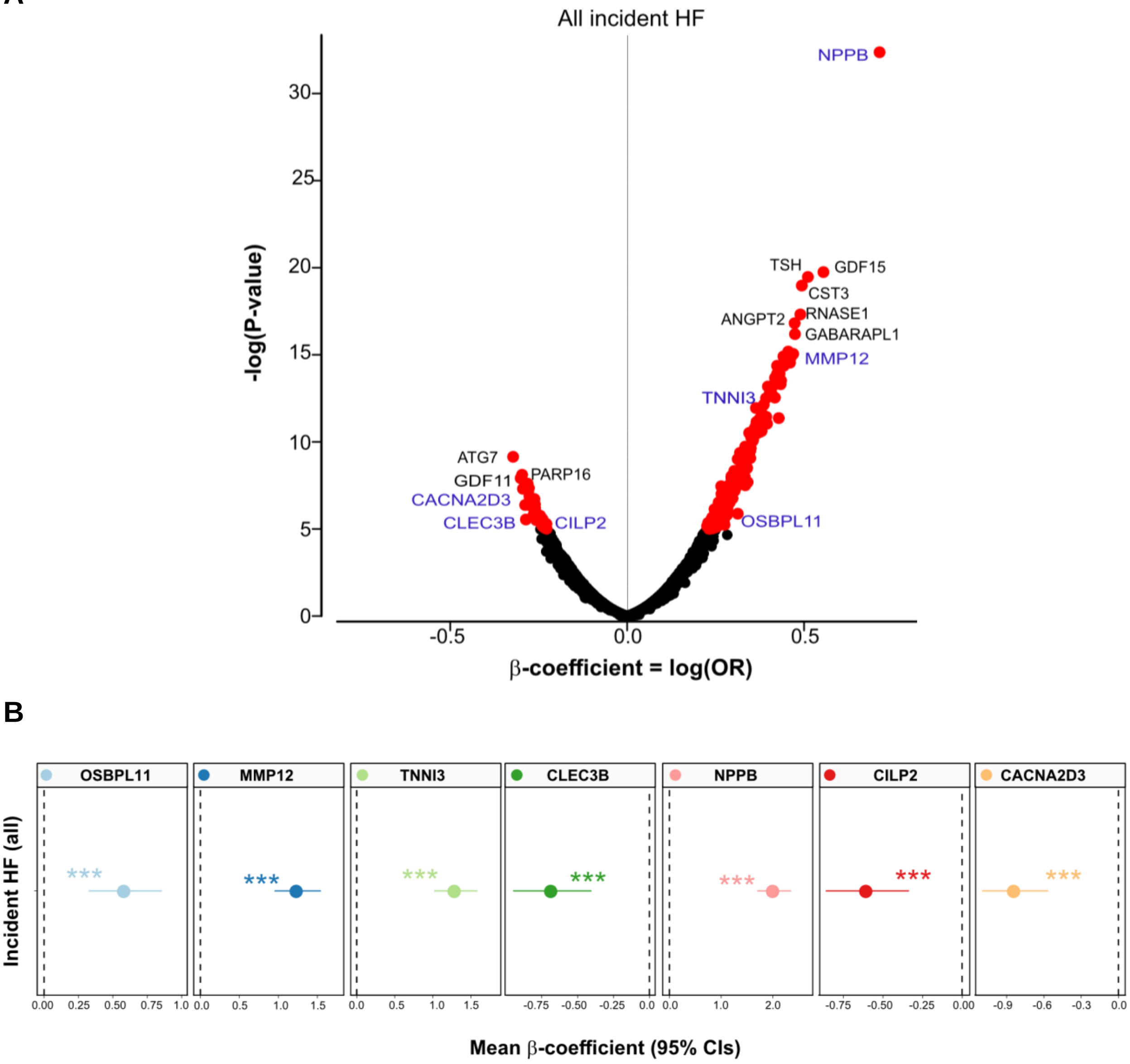


**Figure S5**. **Association of global serum proteins with incident HF**. **A**. a volcano plot of all serum proteins associated with incident HF (all) is shown using age- and sex-adjusted logistic regression analysis and Bonferroni correction for multiple comparisons, with colored (red) data points highlighting study-wide significant associations (P-value < $1\times10^{-5}$, two-sided). Some proteins of interest are highlighted, including those found in the protein predictor for incident HF. The y-axis represents the $-\log_{10}$(P-value) of associations (linear regression), while the x-axis represents the estimate as beta coefficient = log(OR). **B**. highlights the comparison of the top and bottom quintiles of the seven protein predictors associated with incident HF. The data points are the mean estimate of beta in the logistic regression (logOR) using quintiles as continuous predictor variable and the error bars represent 95% CIs. ***(P-value < 0.001, two-sided).

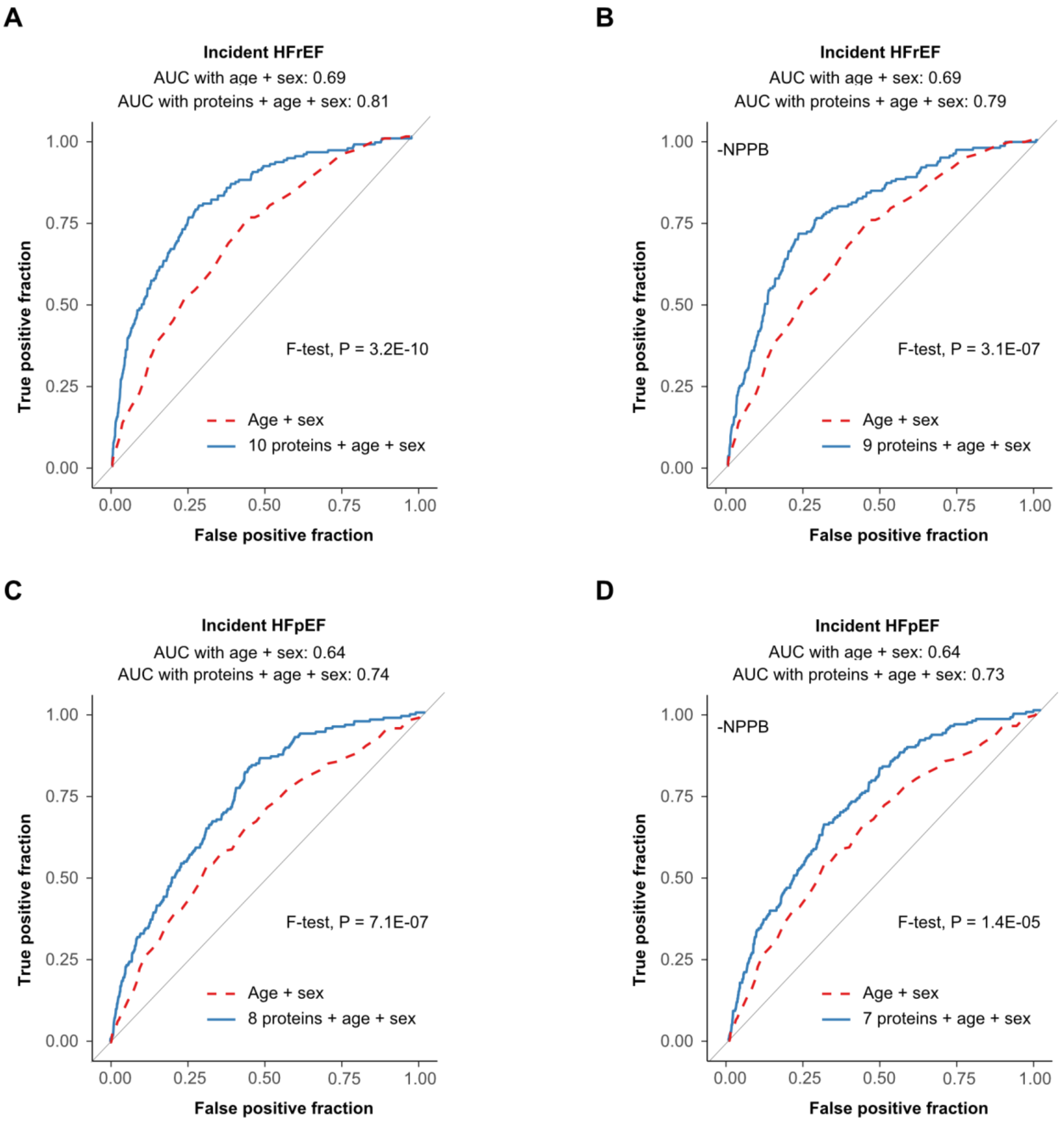


**Figure S6**. **The ability of serum protein predictors to classify incident HFrEF or HFpEF**. A receiver operating characteristic curve (ROC) for the ability of **A**. all 10 protein predictors from Figure 2B to classify incident HFrEF and **B**. when NPPB is excluded from the panel. **C**. shows the ROC curve for all 8 protein predictors from Figure 2B for incident HFpEF, while **D**. has excluded NPPB from the protein panel. The F-tests of equality P-values (two-sided) show significant differences between the demographics (age + sex, red broken curve) ROC curves and the demographics plus proteins (age + sex + proteins, blue solid curve).

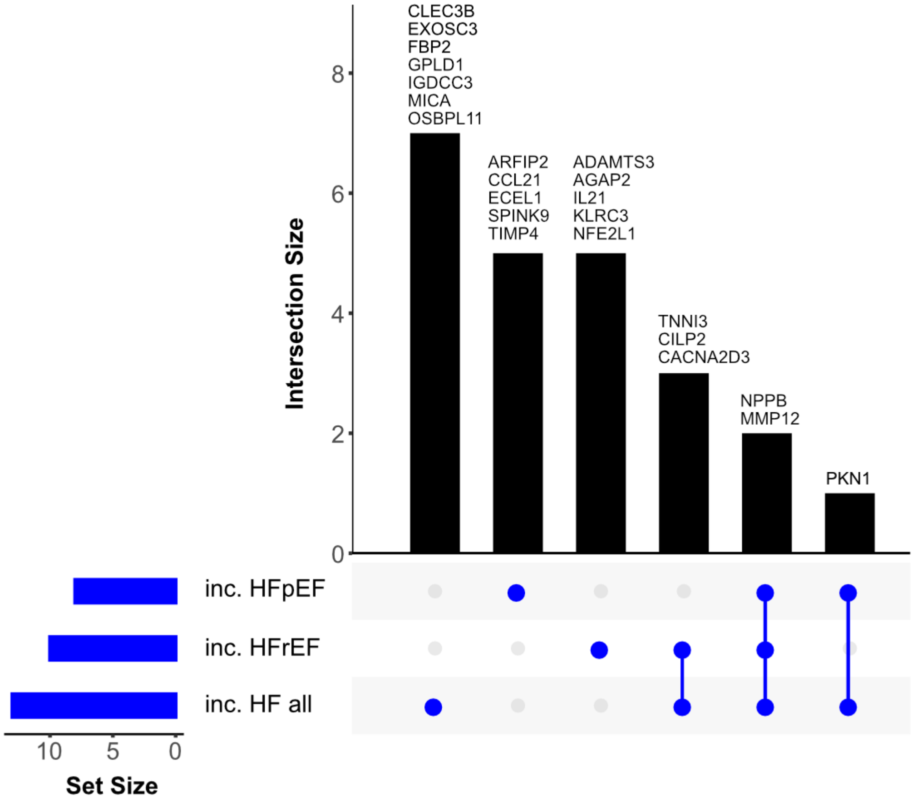


**Figure S7**. **An UpSet plot displaying the overlap between the various protein predictors for incident HF**. The protein predictors that overlap or are unique for all incident HF, incident HFrEF, and/or HFpEF are listed at the top of each bar in the graph.

A

B

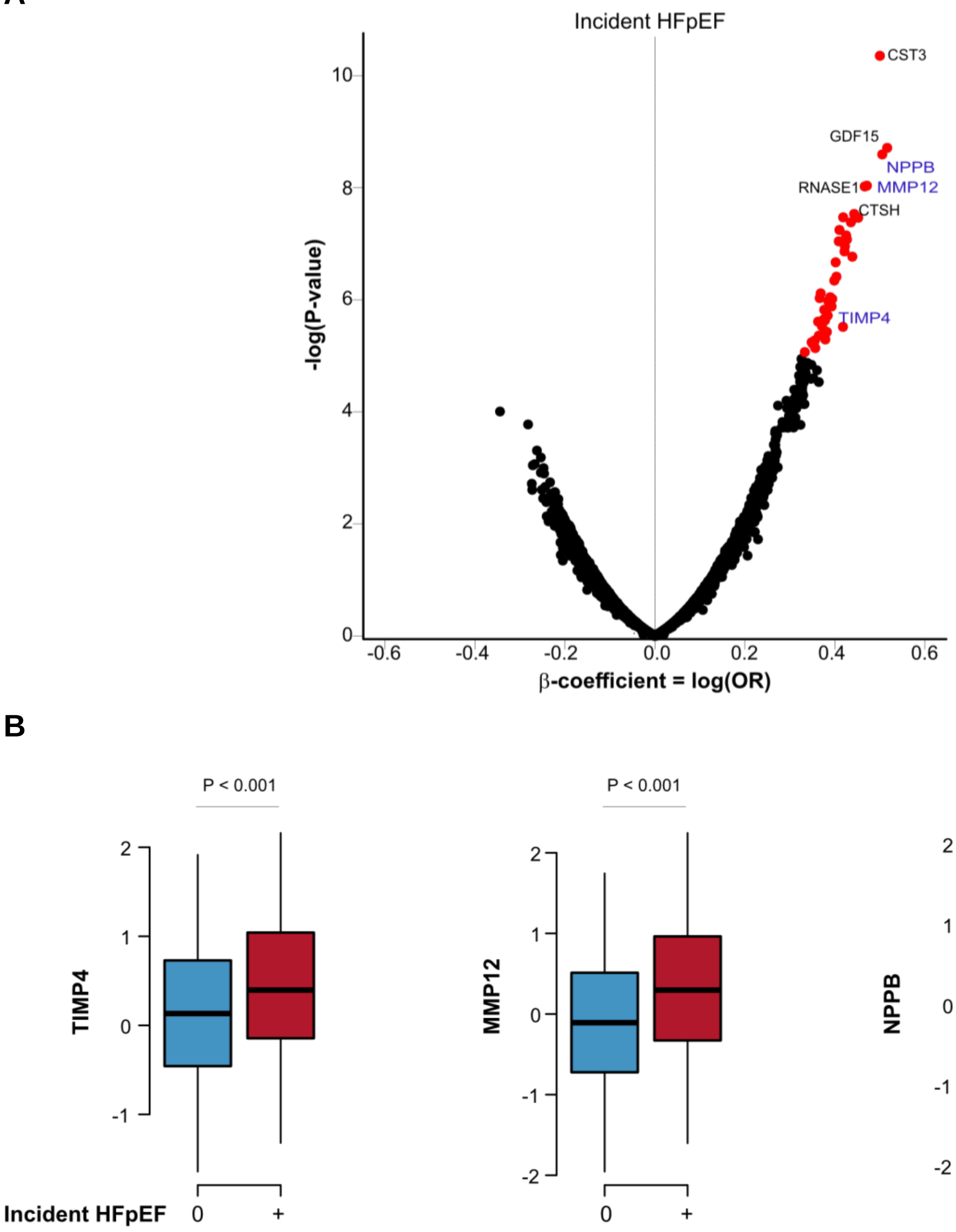


**Figure S8**. **Association of global serum proteins with incident HFpEF**. **A**. a volcano plot of all serum proteins associated with incident HFpEF is shown using age- and sex-adjusted logistic regression analysis and Bonferroni correction for multiple comparisons, with colored (red) data points highlighting study-wide significant associations (P-value < $1 \times 10^{-5}$, two-sided). Some proteins of interest are highlighted, including those found in the protein predictor for incident HFpEF The y-axis represents the $-\log_{10}$(P-value) of associations (linear regression), while the x-axis represents the estimate as beta coefficient = log(OR). **B**. Boxplots depicting the differential expression of three protein predictors in incident HFpEF versus those without HF. All box plots in the figure show median (middle line), 25th, 75th percentile (box) and 5th and 95th percentile (whiskers).

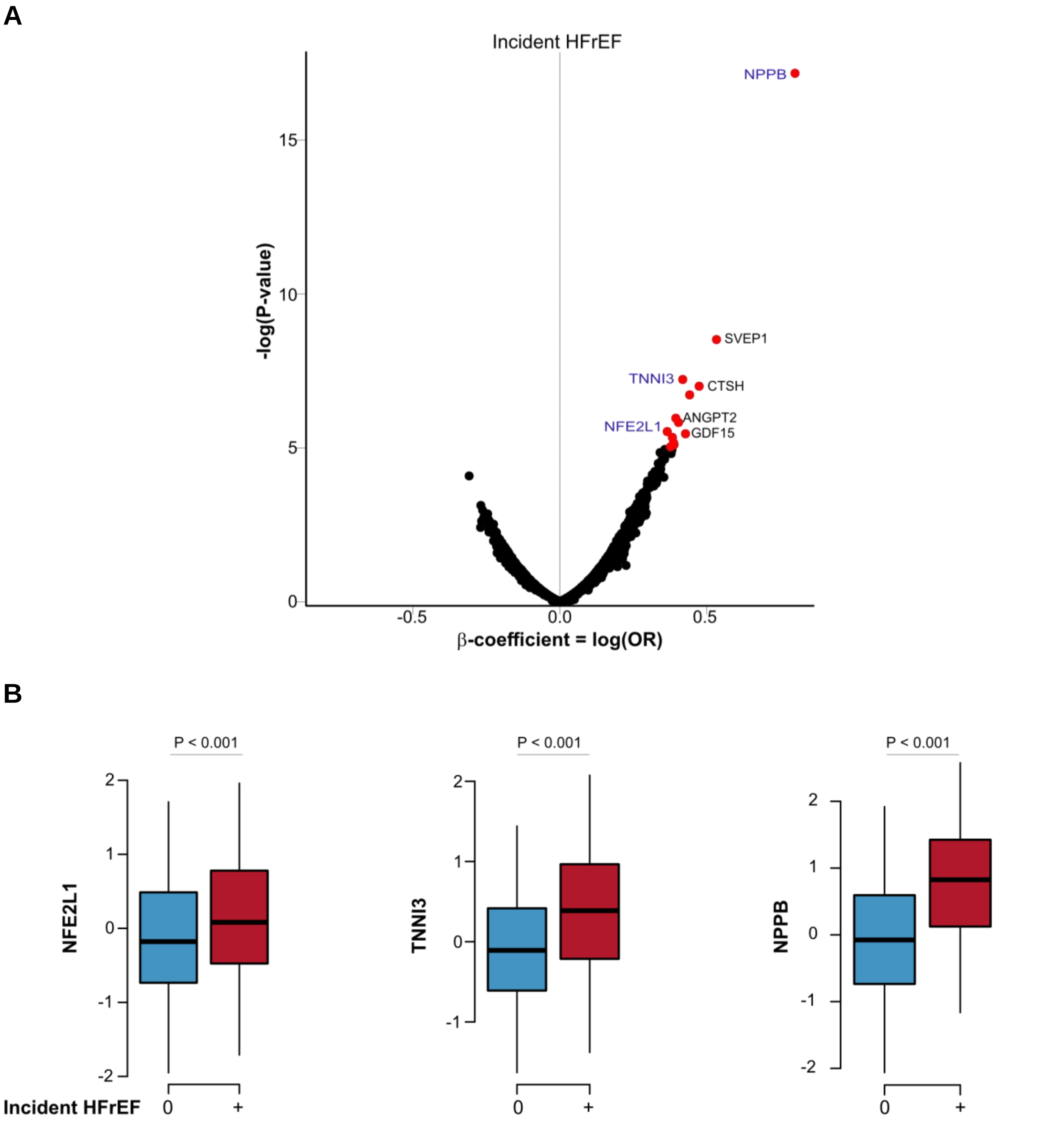


**Figure S9**. **Association of global serum proteins with incident HFrEF**. **A**. a volcano plot of all serum proteins associated with incident HFrEF is shown using age- and sex-adjusted logistic regression analysis and Bonferroni correction for multiple comparisons, with colored (red) data points highlighting study-wide significant associations (P-value < $1\times10^{-5}$, two-sided). Some proteins of interest are highlighted, including those found in the protein predictor for incident HFrEF The y-axis represents the -$\log_{10}$(P-value) of associations (linear regression), while the x-axis represents the estimate as beta coefficient = log(OR). **B**. boxplots depicting the differential expression of three protein predictors in incident HFrEF versus those without HF. All box plots in the figure show median (middle line), 25th, 75th percentile (box) and 5th and 95th percentile (whiskers).

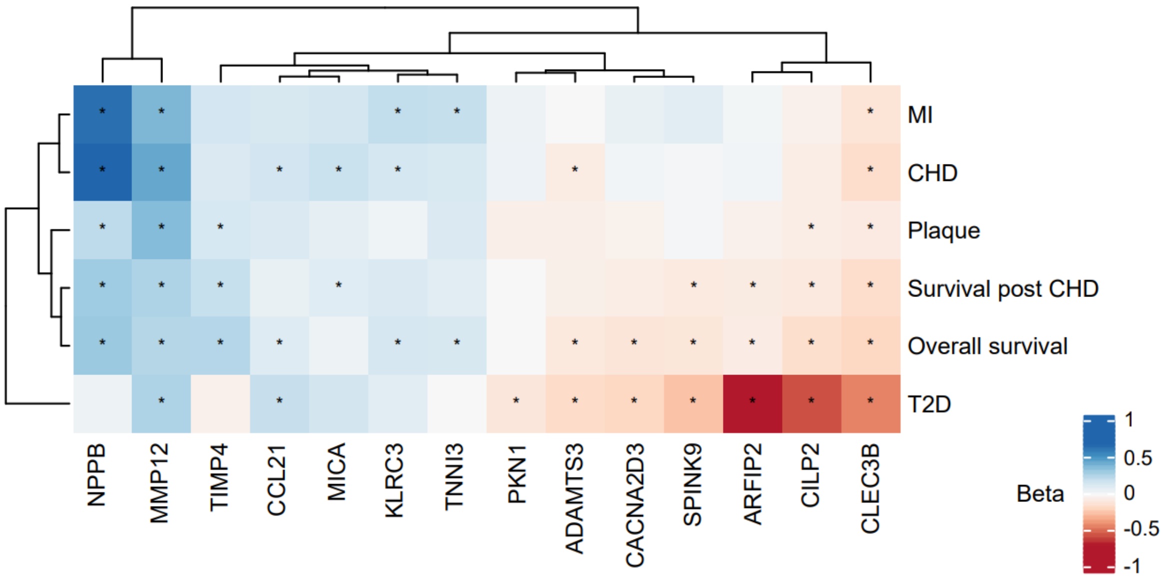


**Figure S10**. **A heatmap depicting the relationship between protein predictors and various outcomes.** The red squares show an inverse relationship between protein predictors and outcome, whereas the blue squares show a direct relationship. After Bonferroni correction for multiple comparisons (Padj < 0.05), the star in the box indicates a significant association between proteins and outcome. Protein predictors that had no statistically significant association with any of the outcome data are not displayed.

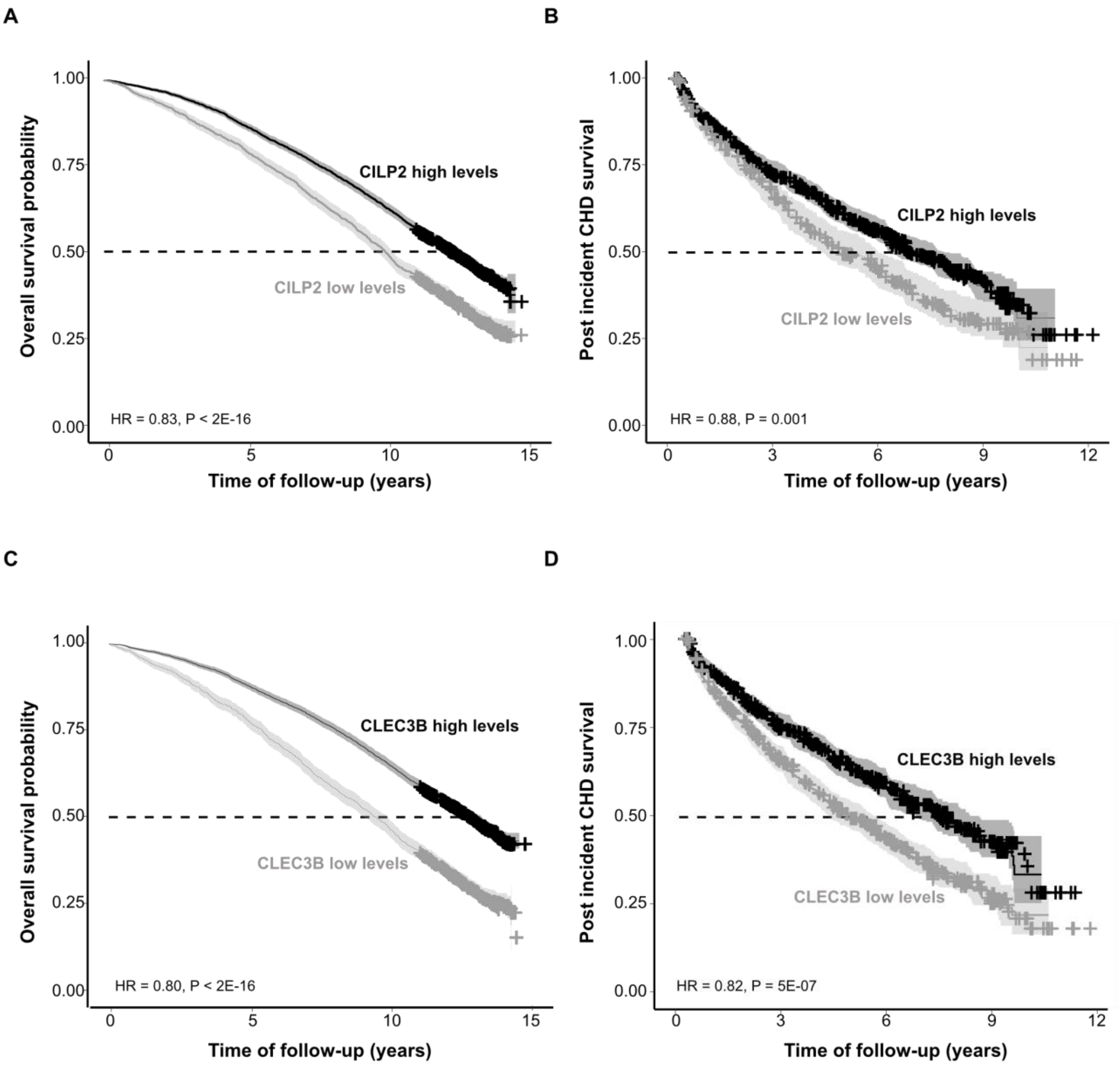


**Figure S11**. **Survival analysis of serum proteins that predict incident HF**. **A**. Kaplan–Meier survival curves showing lower serum levels of CILP2 are significantly associated with a lower overall survival probability (all-cause mortality, n = 2,982 events) as well as **B**. reduced survival post incident CHD (692 events). **C**. Kaplan–Meier curves showing lower serum levels of CLEC3B are associated with increased all-cause mortality, and **D**. reduced survival probability post incident CHD.

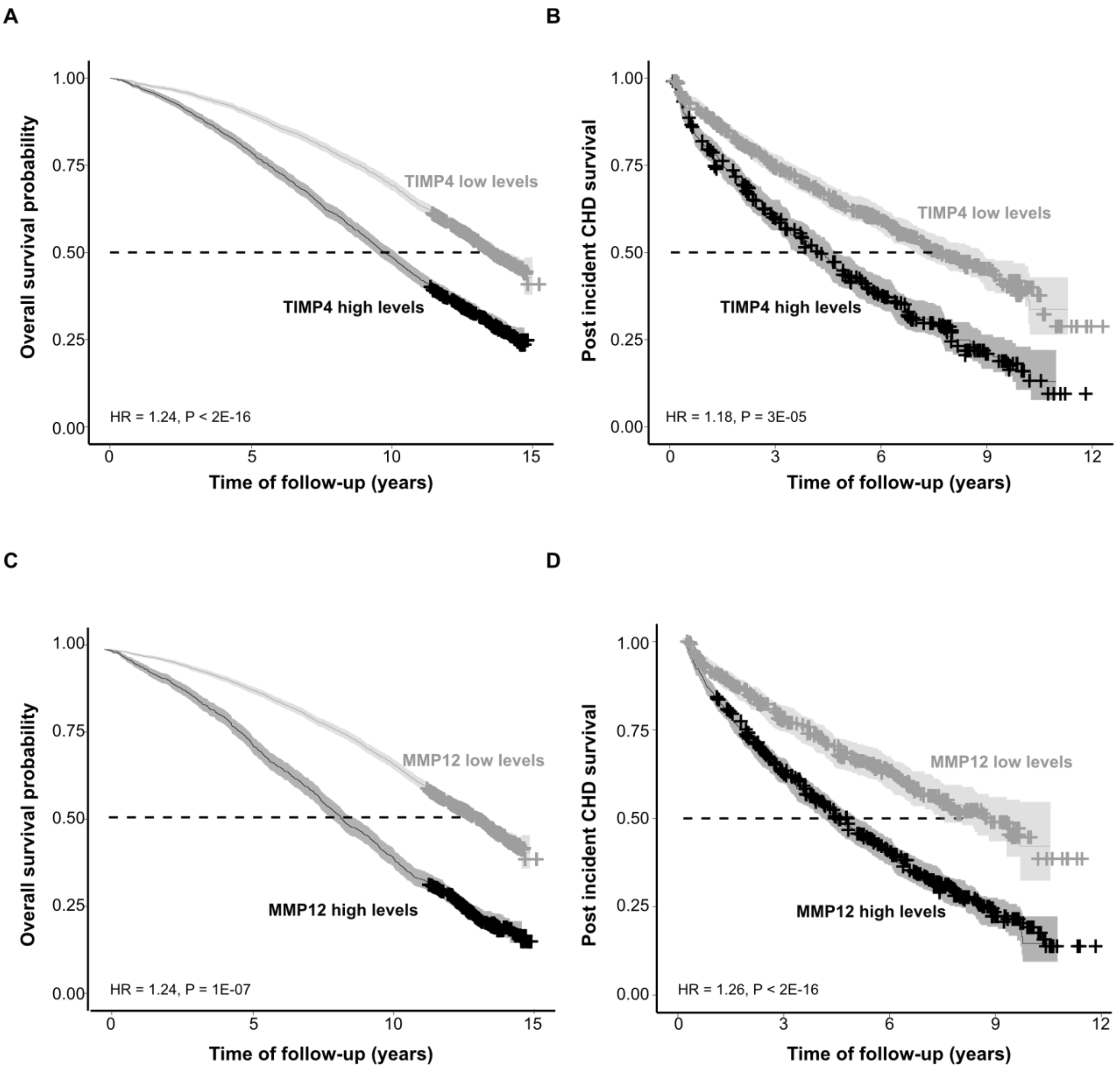


**Figure S12**. **Survival analysis of serum proteins that predict incident HF**. **A**. Kaplan–Meier survival curves showing elevated serum levels of TIMP4 are significantly associated with a lower overall survival probability (all-cause mortality, n = 2,982 events) as well as **B**. reduced survival post incident CHD (692 events). **C**. Kaplan–Meier curves showing higher serum levels of MMP12 are associated with increased rates of all-cause mortality and **D**. reduced survival probability post incident CHD.

**A**

**Proteins that respond to SGLT2 treatment and are linked to incident HF**

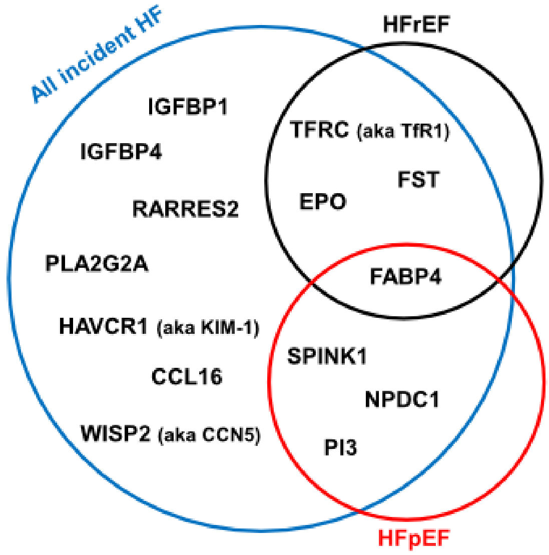


| Protein | Resp. to SGLT2 | All incident HF | | HFrEF | | HFpEF | |
|---|---|---|---|---|---|---|---|
| | | β | P-value | β | P-value | β | P-value |
| CCL16 | ↑ | 0.272 | 2.1E-07 | N/A | NS | 0.277 | 0.0003 |
| EPO | ↑ | 0.334 | 1.9E-10 | 0.395 | 1.1E-06 | N/A | NS |
| FABP4 | ↑ | 0.435 | 3.0E-14 | 0.382 | 3.8E-06 | 0.379 | 1.4E-05 |
| FST | ↑ | 0.193 | 0.0007 | 0.394 | 0.0001 | N/A | NS |
| HAVCR1 | ↓ | 0.260 | 8.3E-07 | N/A | NS | N/A | NS |
| IGFBP1 | ↑ | 0.171 | 0.0018 | N/A | NS | N/A | NS |
| IGFBP4 | ↑ | 0.251 | 2.5E-06 | N/A | NS | N/A | NS |
| NPDC1 | ↑ | 0.350 | 5.3E-11 | N/A | NS | 0.419 | 3.4E-08 |
| PI3 | ↑ | 0.323 | 1.5E-08 | N/A | NS | 0.277 | 0.0008 |
| PLA2G2A | ↑ | 0.245 | 2.6E-06 | N/A | NS | N/A | NS |
| RARRES2 | ↑ | 0.270 | 9.1E-07 | N/A | NS | N/A | NS |
| SPINK1 | ↑ | 0.234 | 6.2E-06 | N/A | NS | 0.280 | 0.0002 |
| TFRC | ↑ | 0.252 | 2.3E-06 | 0.300 | 0.0003 | N/A | NS |
| WISP2 | ↑ | 0.275 | 5.8E-06 | N/A | NS | N/A | NS |

**B**

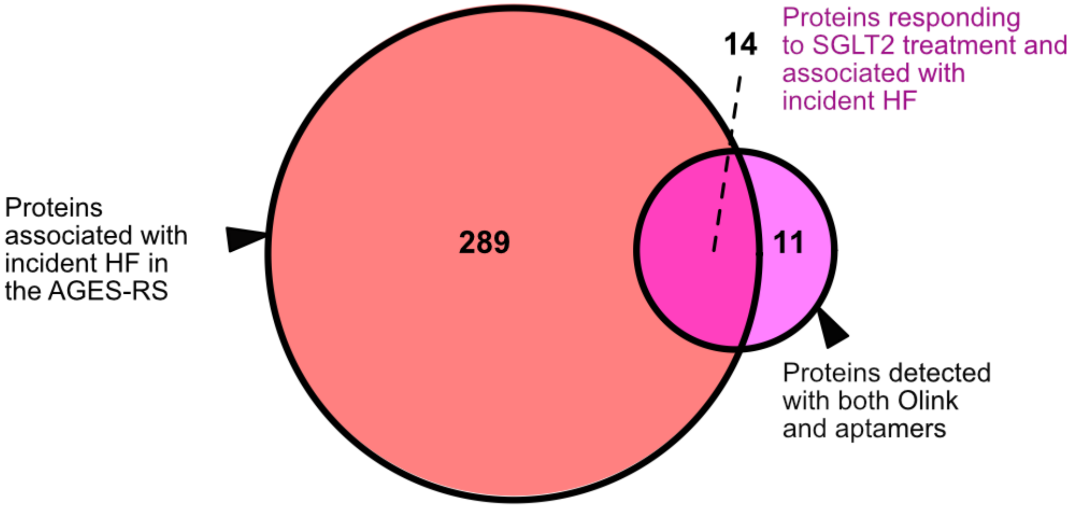


**FET OR = 9.4, P = 1.6E-08**

**Figure S13**. **Proteins that respond to SGLT2 treatment and associated with incident HF**. **A**. All proteins responding to SGLT2 treatment and found associated with incident HF including HFrEF and/or HFpEF. **B**. Enrichment (FET, Fisher exact test) of SGLT2 proteins responders (PMID: 36017745) among proteins associated with incident HF in AGES-RS.
